# Evaluation of variant calling algorithms for wastewater-based epidemiology using mixed populations of SARS-CoV-2 variants in synthetic and wastewater samples

**DOI:** 10.1101/2022.06.06.22275866

**Authors:** Irene Bassano, Vinoy K. Ramachandran, Mohammad S. Khalifa, Chris J. Lilley, Mathew R. Brown, Ronny van Aerle, Hubert Denise, William Rowe, Airey George, Edward Cairns, Claudia Wierzbicki, Natalie D. Pickwell, Myles Wilson, Matthew Carlile, Nadine Holmes, Alexander Payne, Matthew Loose, Terry A. Burke, Steve Paterson, Matthew J. Wade, Jasmine M.S. Grimsley

## Abstract

Wastewater-based epidemiology (WBE) has been used extensively throughout the COVID-19 pandemic to detect and monitor the spread and prevalence of SARS-CoV-2 and its variants. It has proven an excellent, complementary tool to clinical sequencing, supporting the insights gained and helping to make informed public health decisions. Consequently, many groups globally have developed bioinformatics pipelines to analyse sequencing data from wastewater. Accurate calling of mutations is critical in this process and in the assignment of circulating variants, yet, to date, the performance of variant-calling algorithms in wastewater samples has not been investigated. To address this, we compared the performance of six variant callers (VarScan, iVar, GATK, FreeBayes, LoFreq and BCFtools), used widely in bioinformatics pipelines, on 19 synthetic samples with known ratios of three different SARS-CoV-2 variants (Alpha, Beta and Delta), as well as 13 wastewater samples collected in London between the 15–18 December 2021. We used the fundamental parameters of recall (sensitivity) and precision (specificity) to confirm the presence of mutational profiles defining specific variants across the six variant callers.

Our results show that BCFtools, FreeBayes and VarScan found the expected variants with higher precision and recall than GATK or iVar, although the latter identified more expected defining mutations than other callers. LoFreq gave the least reliable results due to the high number of false-positive mutations detected, resulting in lower precision. Similar results were obtained for both the synthetic and wastewater samples.

## Introduction

On March 11^th^, 2020, the World Health Organisation (WHO) declared a global pandemic following the rapid spread of a novel coronavirus, severe acute respiratory syndrome coronavirus 2 (SARS-CoV-2) [1], which causes coronavirus disease 19 (COVID-19). Since then, wastewater-based epidemiology (WBE) has proven a promising tool to detect and monitor SARS-CoV-2, act as a proxy for infections within certain regions/communities and provide an early-warning of disease outbreaks [2]. It has been widely used across the globe to complement conventional clinical surveillance, which is limited in population coverage, capacity or engagement (e.g., self-testing/reporting) [3]. In this regard, it is evident that WBE can be used to monitor disease prevalence in a community, allowing targeted public health measures to be implemented at relative pace and geographical specificity, in combination with other data. Moreover, WBE is non-invasive and less biased than clinical data, making it a valuable molecular surveillance tool [4, 5].

Since the initial outbreak of SARS-CoV-2, several variants of concern (VOCs), variants under investigation or monitoring (VUIs/VUMs) and variants of interest (VOIs) have circulated globally. According to the WHO, as of May 2022, there have been five VOCs (Alpha, Beta, Gamma, Delta and Omicron), eight VOIs (Epsilon, Zeta, Eta, Theta, Iota, Kappa, Lambda, Mu) and two VUIs (B.1.640 and XD) [6]. While VOCs have transmitted worldwide, VUIs are country-specific, with over 200 sub-lineages of the main circulating variants reported by individual countries [7]. In England, the Horizon Scanning Programme, part of the UK Health Security Agency (UKHSA), has been monitoring circulating variants, including VOC, VUI/VUM and VOIs, identified by deep sequencing of a large cohort of Covid-19 positive patients by COG-UK (Covid-19 Genomics UK) [8, 9]. Since the declaration of the pandemic, this totals over 2 million patients in the UK alone [10]. However, the sequencing datasets generated lacked asymptomatic cases and cases not sequenced.

The Environmental Monitoring for Health Protection programme (EMHP), part of the UKHSA, has used reports produced by the Horizon Scanning Programme to monitor the same variants in wastewater collected in England. However, the analysis of SARS-CoV-2 sequences from wastewater samples is more complicated than clinical samples obtained using nasopharyngeal swabs. Discrepancies between clinical and wastewater samples have been observed; in particular, the mixed strain nature of wastewater samples, the more degraded nature of viral genomes and, consequently, the inability to obtain consensus genome sequence for each of the samples analysed. These differences can be accounted for by the nature and characteristics of the samples (wastewater vs clinical) and characteristics impacting the ability to extract and analyse the samples, such as virus titre, which is considerably lower in wastewater samples that, in turn, may affect variant calling such as sample preparation, with additional steps such as centrifugation and filtration methods required to purify the samples from chemicals and other sources of contamination and platform dependent sequencing errors [11-14].

Several bioinformatics pipelines have been developed to specifically detect SARS-CoV-2 sequences and variants in wastewater samples, including nfcore/viralrecon [15] and V-PIPE SARS-CoV-2 [16]. However, most studies have relied heavily on the ARTIC pipeline initially designed with clinical samples in mind, or, as in the case of the EMHP programme in England, an adaptation of this pipeline. Common sequencing pipelines, including ARTIC, involve the removal of low-quality sequencing reads, followed by read mapping and variant calling to define mutations found in the sample (Single Nucleotide Polymorphisms – SNPs – and INsertions/DELetions) [17]. This is performed by highly specialised tools known as variant callers. The ARTIC pipeline for sequence analysis of clinical samples utilises iVar, which relies on the samtools mpileup command as its variant calling function [17]. While this has been well documented in clinical studies, very little is known about its performance for wastewater samples. To address this knowledge gap, the EMHP programme in England adapted this protocol, using VarScan as an alternative to iVar, delivering significantly improved results when applied to wastewater samples [18, 19]. Before screening for sequence changes, VarScan uses the BAM alignment file as the input to score each of the reads produced during sequencing. If reads are found to align to multiple locations and/or are of low quality, they are automatically discarded. For the remaining reads, SNPs and Indels are compiled for each of the locations across the viral genome and validated depending on factors such as the overall coverage, the number of reads across the site of the mutation and base quality, among others.

Several genomic studies have compared and highlighted the impact that variant caller choice has on the analysis pipeline, including iVar [17], GATK [20], LoFreq [21], FreeBayes [22] and BCFtools [23]. FreeBayes is a haplotype-based variant caller where variants are called based on the sequences of reads aligned to a particular target rather than the specific alignment. One of the main advantages of this method is that it bypasses the problem of identical sequences that might align to multiple locations. On the other hand, GATK uses HaplotypeCaller as a tool to call germline SNPs and Indels via local re-assembly of haplotypes [22]. More specifically, it assembles and realigns reads to their most likely haplotypes. Comparison with the reference of choice is used to calculate the likelihood of each possible genotype and call possible variants. LoFreq is a high-quality and highly sensitive tool to detect variants in heterogeneous samples, such as tumour samples [21]. It was developed under the assumption that it is hard to distinguish true variants from sequencing errors. In this regard, LoFreq is a very robust and sensitive variant caller that uses base-called quality values to call variants accurately. It differs from other callers as it can find SNPs and Indels at a frequency below the average sequencing error. As such, it is not ideal for low-coverage genomes. BCFtools is a collection of several commands, among which *call* is used for SNP/indel calling [23]. It generates the *mpileup* from the BAM alignment reads and then computes the variant calling. This step is the same as VarScan, which generates *mpileup* using SAMtools. iVar uses the output of the SAMtools *mpileup* command to call variants as VarScan and BCFtools; however, it is not adapted for use in mixed strain samples, such as those derived from wastewater where mixed populations are found in the same sample [17]. [24, 25]. Indeed, it is globally acknowledged that the detection of defining SNPs and Indels allows the assignment of VOCs, VUIs, VUMs and VOIs to wastewater samples, thus their accurate identification is paramount for variant detection in the context of WBE.

In this manuscript, we evaluated the performance of six different variant callers and their ability to detect SNPs and indels in samples containing a mixture of synthetic SARS-CoV-2 control variants, as well as wastewater samples collected across Greater London during the pandemic.

## Methods

### Sample library preparation and sequencing

The synthetic SARS-CoV-2 control variant dataset contained samples that consisted of a mix of three variant genomes: Alpha (Control 15), Beta (Control 16) and Delta (Control 23) synthesised by TWIST Biosciences, USA (Table 1). For each sample in the synthetic dataset, three of these genomes were mixed in different ratios up to a total concentration of 200 genome copies per □L, ranging from 0 to 100% of each synthetic genome, in quadruplicates. The mutation profile of each of the synthetic genomes is provided in Supplementary Table 1. Wastewater samples were collected between the 15–18 December 2021 in eight locations across the city of London, UK. Most of these samples were found to be positive for both the Delta and Omicron lineages using the bioinformatics pipeline developed by EMHP (data not shown). Wastewater samples were clarified, concentrated and RNA extracted according to the Quantification of SARS-CoV-2 in Wastewater General Protocol V1.0 (https://www.cefas.co.uk/media/offhscr0/generic-protocol-v1.pdf). Sequencing libraries (tiled amplicons) were generated using the EasySeq™ SARS-CoV-2 WGS Library Prep Kit (Nimagen, The Netherlands) and the Nimagen V3 (wastewater samples) and V4 (synthetic samples) primer schemes, following the Wastewater Sequencing using the EasySeq™ RC-PCR SARS-CoV-2 (Nimagen) V2.0 protocol [26]. The libraries were sequenced on an Illumina NovaSeq 6000 (2x 150 bp) at the University of Liverpool sequencing centre (synthetic samples) or an Illumina NextSeq 500 (2x 150 bp) at the University of Nottingham sequencing centre (wastewater samples).

**Table 1.** List of synthetic genomes by Twist Bioscience used for the comparison test. Defining mutation patters were taken from https://github.com/phe-genomics/variant_definitions.

| Twist Part No | GISAID ID | GISAID Name | PANGO lineage | WHO Label | Defining mutation | Total mutations in Twist genome | SNPs | Indels | MNVs |
| --- | --- | --- | --- | --- | --- | --- | --- | --- | --- |
| 103909 | EPI_ISL_601443 | England/MILK9E05B3/2020 | B.1.1.7 | Alpha | 15 | 28 | 22 | 4 | 2 |
| 104043 | EPI_ISL_678597 | South Africa/KRISPEC-K005299/2020 | B.1.351 | Beta | 15 | 25 | 23 | 2 | 0 |
| 104533 | EPI_ISL_1544014 | India/MH-NCCS P1162000182735/2021 | B.1.617.2 | Delta | 13 | 37 | 30 | 7 | 0 |
| 104044 | EPI_ISL_792683 | Japan/IC-0564/2021 | P.1 | Gamma | 23 | 24 | 21 | 3 | 0 |
| 105204 | EPI_ISL_6841980 | Hong Kong/HKU-211129-001/2021 | B.1.1.529 BA.1 | Omicron | 17 | 59 | 45 | 14 | 0 |
| 105345 | EPI_ISL_7190366 | Australia/QLD2568/202 | B.1.1.529 BA.2 | Omicron | 20 | 57 | 48 | 9 | 0 |
| 105346 | EPI_ISL_7718520 | England/MILK-2DF642C/2021 | B.1.1.529 BA.2 | Omicron | 20 | 58 | 48 | 9 | 0 |

### Read pre-processing, mapping, primer trimming and variant calling

The ARTIC pipeline (ncov2019-artic-nf; Illumina workflow) [27] was used to process the raw Illumina reads. Briefly, amplicon reads were pre-processed using Trim Galore v0.6.5 [28], mapped to the reference SARS-CoV-2 genome (ENA GenBank Accession MN908947.3, NCBI NC_045512.2) using BWA v0.7.17 [29], followed by primer trimming using iVar v1.3 and bed files containing the genome positions of the primers used to generate the amplicons (Nimagen V3 and V4 primer schemes for wastewater and synthetic samples, respectively). The resulting BAM files were sorted and subsequently indexed using SAMtools v1.13 [23] before analysis with six different variant callers; iVar v1.3.1, LoFreq v2.1.3.1, BCFtools v1.13, GATK Haplotypecaller v3.8, VarScan v2.4.4 and FreeBayes v0.9.21. To avoid introducing biases across the variant callers, only parameters common to those available from VarScan were chosen. VarScan is the caller with the least number of parameters as it allows to only choose from min-coverage (Minimum read depth at a position to make a call), min-reads2 (Minimum supporting reads at a position to call variants), min-avg-qual (Minimum base quality at a position to count a read), min-var-freq (Minimum variant allele frequency threshold) and p-value (p-value threshold for calling variants). It should be also noted that it is practically impossible to test all the parameters from all the callers and leaving these in default is a preferred choice when performing comparison studies [30-32].

The ARTIC pipeline outputs a list of mutations (SNPs and indels) detected for each variant using iVar, but this was re-run separately after matching the common parameters across the various callers being investigated. All parameters are described in Supplementary file 1. The sorted, indexed and primer-trimmed BAM files were used directly to run variant calling with FreeBayes, iVar, BCFtools and VarScan, while LoFreq and GATK Haplotypecaller required first pre-processing of these BAM files. Since LoFreq required indel quality information in the BAM file to process indel calls, we used the LoFreq command *indelqual* to insert quality score for each indel, based on the dindel algorithm [33]. GATK Haplotypecaller, requires reads to be grouped (using *AddOrReplaceReadGroups* from Picard) and duplicates (*MarkDuplicatesSpark* from GATK) were marked before variant calling. All the variant callers generated outputs in the variant call format (vcf) files except iVar, which reported outputs as tsv (tab-separated values) files. A python script (ivar_variants_to_vcf.py) was used to convert the tsv file to vcf format [15]. A python script (<u>ivar_variants_to_vcf.py</u>) was used to convert the tsv file to vcf format [34].

### VCF file processing, analysis, and statistical methods

QuasiModo is a tool that evaluates the results of strain resolved analyses on mixed strain samples including variant calling and genome assembly [35]. It does this by taking vcf files generated from the different variant callers and two genomic reference files, the first being the reference against which samples were mapped in the BAM file-generating process, and the second reference being a ground-truth genome known to be found in the mixed strain samples. We therefore evaluated the performance of the different variant callers by comparing lists of mutations identified by each of the variant callers to a second reference genome (for ground truthing). The reference SARS-CoV-2 genome sequence was downloaded from NCBI Genbank (Accession No. MN908947.3) and the SARS-CoV-2 variant genomes were obtained from GISAID: Alpha (EPI_ISL_601443), Beta (EPI_ISL_678597), Delta (EPI_ISL_1544014), Omicron-England (EPI_ISL_7718520), Omicron-Hong Kong (EPI_ISL_6841980), Omicron-Australia (EPI_ISL_7190366) and Gamma (EPI_ISL_792683). Briefly, sequences from each sample were aligned to the reference genomes using MUMmer4 [27] to identify SNPs and indels that are present in the ground-truth genome and each variant call was then categorised as either a true positive (TP), a false positive (FP), or a false negative (FN). A true positive is defined as one that was found by the variant caller being tested in both the sample and the reference. A true negative is a lack of a mutation detected by the variant caller where there is no mutation present in the reference file. A false positive is a mutation reported by the variant caller but not present in the original reference, while a false negative is a mutation not detected by the variant caller, but that is found in the reference [35-38].

These values are used to calculate the recall and precision, also known as sensitivity and specificity, respectively:

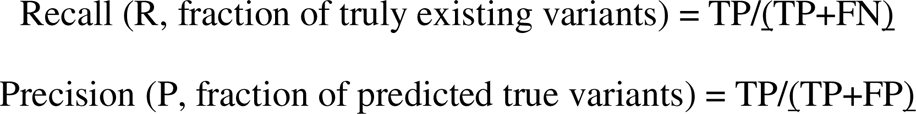

In addition, once recall and precision are calculated, a ratio of the two can be derived, known as the F1 score [35]:

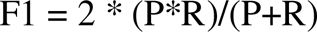

For the synthetic control samples, 455 vcf files (generated for 19 synthetic samples (quadruplicates) from six variant callers, with one failed replicate for GATK) were analysed using the MN908947.3 reference file as the mapping genome and each of the SARS-CoV-2 variant reference genomes (Alpha, Beta, Delta, and Gamma). Similarly, for wastewater samples, we generated 77 vcf files from 13 unique samples among six variant callers, with one failed sample for GATK), using the MN908947.3 reference and each of the SARS-CoV-2 variant genomes (Alpha, Beta and Delta, Omicron (Hong Kong), Omicron (Australia) and Omicron (England) and Gamma; Table 1). The Gamma (P1) variant reference file (Table 1) served as a negative control in our bioinformatics analysis, as it was not included in the synthetic mixtures nor found in the wastewater samples. We adapted the method described by Deng et al. [35] to generate a table with calculated values for each of the vcf files, from which recall, and precision were plotted using R v4.1.3 and ggplot2 [39]. Output from all the vcf files was used by *vcfstats* from the vcflib package [40] to generate variant statistics for each of the vcf files. *vcfstats* generates a two-column output for each vcf file, with counts for SNPs, MNPs (multiple nucleotide polymorphisms), Indels and various other parameters. The number of SNPs, Indels and MNPs for each vcf file were plotted using RStudio, ggplot2 package [39]. Frequencies of the defining mutations for each of the variant genomes were extracted from the vcf files using BCFtools [23] and plotted using Rstudio, ggplot2 [39].

To test whether the distribution of precision, recall and F1 scores for each variant caller was significantly different from another, we applied the Kruskal-Wallis one-way analysis of variance test using the python scipy package v1.9.1 (<u>SciPy</u>). Following this, a post-hoc Dunn test using scikit-posthocs package v0.7.0 (<u>scikit-posthocs · PyPI</u>) was performed to evaluate the pairwise differences between callers. These tests were performed separately on the synthetic samples, the wastewater samples and both sets of samples together, for each score. Each set of quadruplicate synthetic samples were aggregated by median score before applying the relevant tests.

## Results

### Sensitivity and specificity of six variant callers across mixed synthetic genome samples

We ran VarScan, GATK, iVar, FreeBayes, LoFreq and BCFtools across 19 mixed ratio synthetic samples, in quadruplicates. Basic sequencing statistics for all the samples calculating recall, precision and F1 score values are summarised in Supplementary Figure 7A.

We calculated recall and precision for each variant within the samples and plotted these separately for each variant caller (Figure 1A-D, Supplementary Figure 1A-D), by also highlighting the percentage of each variant in the mixed sample as described in Table 2. Given that all four replicates yielded very similar results (reliable technical replicates), we run the median of these for all the synthetic samples plots. As shown in Supplementary Figure 2, we picked three samples which only contained 100% of one specific variant in the mix, namely Alpha, Beta or Delta, (shown in Table 2 marked by *) to show the validity of the replicates. They all had indeed a similar distribution of the 4 replicates.

**Figure 1.**
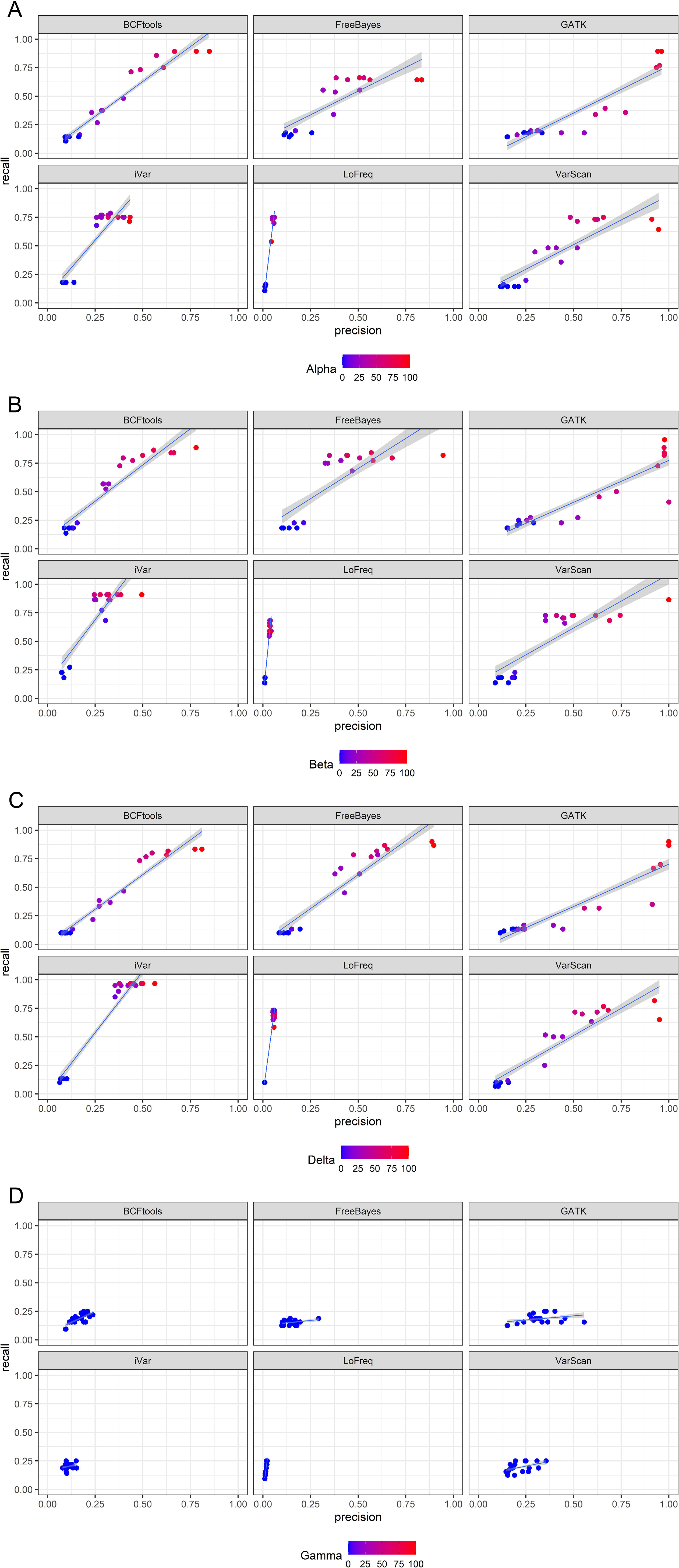
**A-D** Point plots of precision vs recall for synthetic samples, grouped and faceted by variant caller, coloured by percentage present in the mix (dark blue, 0%, bright red, 100%). A linear regression for each plot is also present. A. is comparing the synthetic samples to the alpha variant reference, B. Beta variant reference, C. for Delta variant reference and D. for Gamma variant reference. As there was no mixed ratio for the Gamma variant which we have used as a negative control, no gradient was applied. Except for LoFreq and iVar, all the callers show a high precision and recall, and this is proportional to the percentage of the variant in the mix: indeed, samples with a high percentage of a variant (e.g., close to 100%, in red) tend to have a higher precision and recall, compared to those samples that have a lower percentage of the variant being plotted (e.g., closer to 0%, in blue).

**Table 2.** Mixed synthetic samples used to compare the six variant callers. The samples with * were also used for comparison of technical replication.

| No | Sample mix names | Synthetic (variant) genomes in the sample | Synthetic genome ratio (%) in the mix |
| --- | --- | --- | --- |
| 1* | 100_Alpha\|0_Beta\|0_Delta | Alpha | 100 / 0 / 0 |
| 2* | 0_Alpha\|100_Beta\|0_Delta | Beta | 0 / 100 / 0 |
| 3* | 0_Alpha\|0_Beta\|100_Delta | Delta | 0 / 0 / 100 |
| 4 | 25_Alpha\|75_Beta\|0_Delta | Alpha / Beta | 25 / 75 / 0 |
| 5 | 50_Alpha\|50_Beta\|0_Delta | Alpha / Beta | 50 / 50 / 0 |
| 6 | 75_Alpha\|25_Beta\|0_Delta | Alpha / Beta | 75 / 25 / 0 |
| 7 | 95_Alpha\|5_Beta\|0_Delta | Alpha / Beta | 95 / 5 / 0 |
| 8 | 10_Alpha\|70_Beta\|20_Delta | Alpha / Beta / Delta | 10 / 70 / 20 |
| 9 | 20_Alpha\|70_Beta\|10_Delta | Alpha / Beta / Delta | 20 / 70 / 10 |
| 10 | 25_Alpha\|25_Beta\|50_Delta | Alpha / Beta / Delta | 25 / 25 / 50 |
| 11 | 25_Alpha\|50_Beta\|25_Delta | Alpha / Beta / Delta | 25 / 50 / 25 |
| 12 | 50_Alpha\|25_Beta\|25_Delta | Alpha / Beta / Delta | 50 / 25 / 25 |
| 13 | 50_Alpha\|0_Beta\|50_Delta | Alpha / Delta | 50 / 0 / 50 |
| 14 | 25_Alpha\|0_Beta\|75_Delta | Alpha / Delta | 25 / 0 / 75 |
| 15 | 75_Alpha\|0_Beta\|25_Delta | Alpha / Delta | 75 / 0 / 25 |
| 16 | 0_Alpha\|25_Beta\|75_Delta | Beta / Delta | 0 / 25 / 75 |
| 17 | 0_Alpha\|5_Beta\|95_Delta | Beta / Delta | 0 / 5 / 95 |
| 18 | 0_Alpha\|50_Beta\|50_Delta | Beta / Delta | 0 / 50 / 50 |
| 19 | 0_Alpha\|75_Beta\|25_Delta | Beta / Delta | 0 / 75 / 25 |

Our results show that all the variant callers correctly identified each SARS-CoV-2 variant in the synthetic mixes. At the time of writing, the tools we used to evaluate the presence of variants in a mixed sample via evaluation of their mutational profile could not be applied for mixed samples containing more than two variants or strains; therefore, we investigated the correct identification of the percentage by analysing the variants independently rather than confirming that all variants were found simultaneously in the same sample. Our results suggest that in general, the greater the proportion of a variant (close to 100%), the greater the chance it was called correctly (Figure 1). Indeed, VarScan, BCFtools and FreeBayes correctly called the increased ratio of Alpha compared to the remaining variants in the mix, while iVar and LoFreq had a trend line where the increased concentration of the variants could not be observed as clearly as in the other callers, showing instead a lower precision and for the latter also a low recall (Supplementary Figure 1A-C, 7A). As expected, our negative control P1 (Gamma) (Figure 1D) did not yield any significant results, with all samples having a very low precision and recall for every caller assessed. Specifically, we observed that samples with a ratio close to zero, thus with low concentration of a variant, tended to cluster together with low recall and low precision. Those with higher ratio for a variant, and therefore with more mutations to be detected across several variants, are distributed across the plot to reflect the increased recall, and for some callers, higher precision. Based on this initial observation, BCFtools, VarScan and Freebayes had the highest precision, followed by iVar and GATK. In addition, iVar had the highest recall for each of the variants being assessed independently, via count of their TPs, FPs and FNs compared to the other callers. This was supported by a Dunn’s test to compare the synthetic samples’ precision, recall and F1 scores of each variant caller. It confirmed that the differences in precision and F1 scores for LoFreq were significantly different to the other callers (p < 0.01) and iVar performed best for recall and the Dunn’s test again confirmed this as statistically significant (p < 0.01, Supplementary Figure 7A).

### Sensitivity and specificity of six variant callers across wastewater samples from London

We used wastewater samples collected between the 15–18 December 2021 from Greater London to assess whether the variant callers could identify the mutations with similar precision and recall as observed with the synthetic samples. Table 3 shows the list of the samples, dates and predicted variants known to be found in those samples and Supplementary Figure 7B shows basic sequencing statistics. Samples were predicted to contain a mix of the Omicron and/or Delta, AY.4.2 variants definitions (EMHP analysis based on PHE variant definition, data not shown). Given the genome similarity between the Delta and AY.4.2 variants, we only carried out our analysis on the Delta variant mutations. Figure 2A-B shows that the variant callers recognised mutations that could be identified as Delta variant for some of the samples, which indeed show a higher precision and recall compared to others, while in Supplementary Figure 2A-B-D we show that Alpha, Beta or Gamma variants are not detected, as expected, since these were not expected to be found in the samples, compared to Delta which shows to be found in some of the samples (Supplementary Figure 2C). Indeed, when testing for Delta variant presence, we noticed a slight increase in the precision and recall for some of those samples, which suggests that those did contain SNPs and/or Indels that could be identified as being part of the Delta variant, namely S50 and S296, although the latter was not called by GATK. Consistent with the data in Table 3, some of the samples were found to not contain a Delta variant, such as sample S43, which indeed showed a low precision and recall for all the callers, while other samples with slightly higher values reflected that did contain a mix of both Omicron and Delta (samples S58, S292, S63). As shown for the synthetic samples, LoFreq was the only variant caller that called with the lowest precision for all the samples analysed, followed by iVar, while recall values for LoFreq were sparser, yet higher than the other callers.

**Figure 2.**
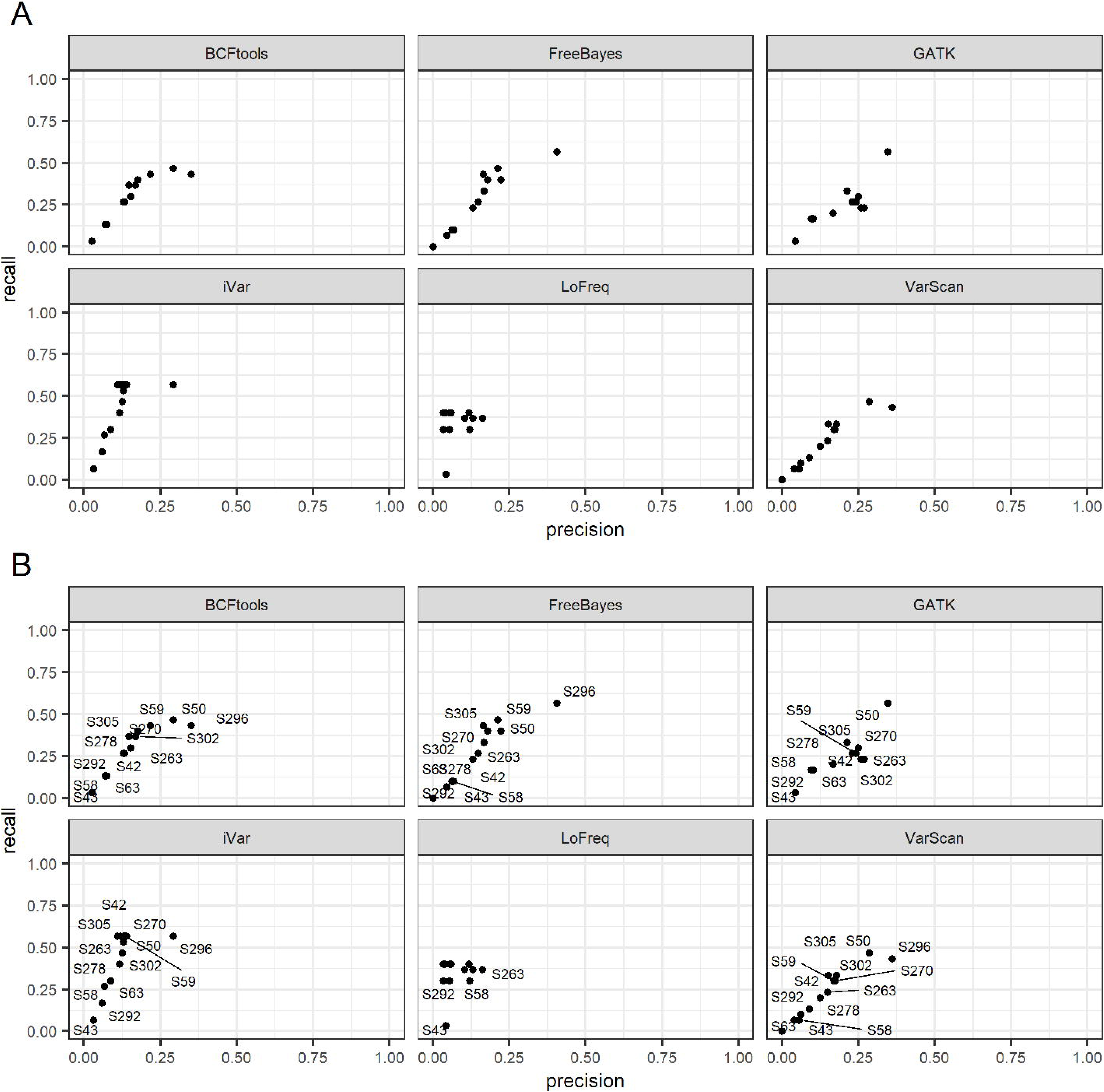
**A-B** Point plots of precision vs recall for wastewater samples for the Delta variant, faceted by variant caller. A. Comparison of real wastewater samples to the Delta variant reference and B. with labelled samples to better identify which samples had low recall and low precisions (thus, not containing any Delta variant). As shown in Figure 4C, some samples such as S296 and S50 are those containing a Delta variant in the mix, compared to S43 seen to be negative for all the variant callers. As shown previously for the synthetic data, LoFreq has the lowest recall and precision.

**Table 3.** List of wastewater samples collected across London between the 15^th^ and 18^th^ December 2021. Samples in bold were also used to generate UpSet plots for Figure 5A-F.

| No | Wastewater Samples | Sample Code | Sample Collection date | Variants detected by GHP |
| --- | --- | --- | --- | --- |
| 1 | London 1 | S59 | 15/12/2021 | Delta / Omicron |
| **2** | **London 2** | **S42** | 16/12/2021 | Delta / Omicron |
| **3** | **London 3** | **S63** | 16/12/2021 | Delta / Omicron |
| 4 | London 4 | S50 | 16/12/2021 | Delta / Omicron |
| **5** | **London 5** | **S58** | 16/12/2021 | Delta / Omicron |
| **6** | **London 6** | **S43** | 16/12/2021 | Omicron |
| **7** | **London 7** | **S296** | 18/12/2021 | Delta |
| **8** | **London 8** | **S263** | 18/12/2021 | Delta / Omicron |
| 9 | London 9 | S302 | 18/12/2021 | Delta / Omicron |
| **10** | **London 10** | **S292** | 18/12/2021 | Delta / Omicron |
| 11 | London 11 | S278 | 18/12/2021 | Omicron |
| 12 | London 12 | S305 | 18/12/2021 | Omicron / AY.4.2 |
| 13 | London 13 | S270 | 18/12/2021 | Omicron / AY.4.2 |

Similarly, we tested the wastewater samples for the presence of the Omicron lineage (Figure 3A-C). We used three different references representative of this variant, namely, England, Hong Kong, and Australia. As shown in Figure 3A-C and Supplementary Figure 4A-C, all three variants under analysis were found in our wastewater samples and at a higher level than the one at which the Delta variant was detected for specific samples expected to have either or both two variants. The degree to which each variant caller recognises the mutations varied, with LoFreq again returning the lowest recall and precision values compared to the other callers. This was consistent with the results obtained with the synthetic data. Based on the predicted detection indicated in Table 3, the two samples identified to contain a Delta based mutational profile as described above (S50 and S296), have now a low precision and recall when tested against any of the Omicron lineages, suggesting that in those samples we can predict to find a Delta variant rather than an Omicron. This was confirmed consistently for all the callers, although we also observed again that LoFreq did have low values as shown in the other plots and that GATK did not call S296. As for the synthetic samples, a Dunn’s test of the pairwise scores confirmed that in terms of precision, GATK, VarScan and Freebayes were not significantly different from one other. However, the Dunn’s test on recall showed iVar to be stochastically dominant. For the combined F1 score, the same test showed that only LoFreq was significantly different from each of the other tools (p < 0.01, Supplementary Figure 7B).

**Figure 3.**
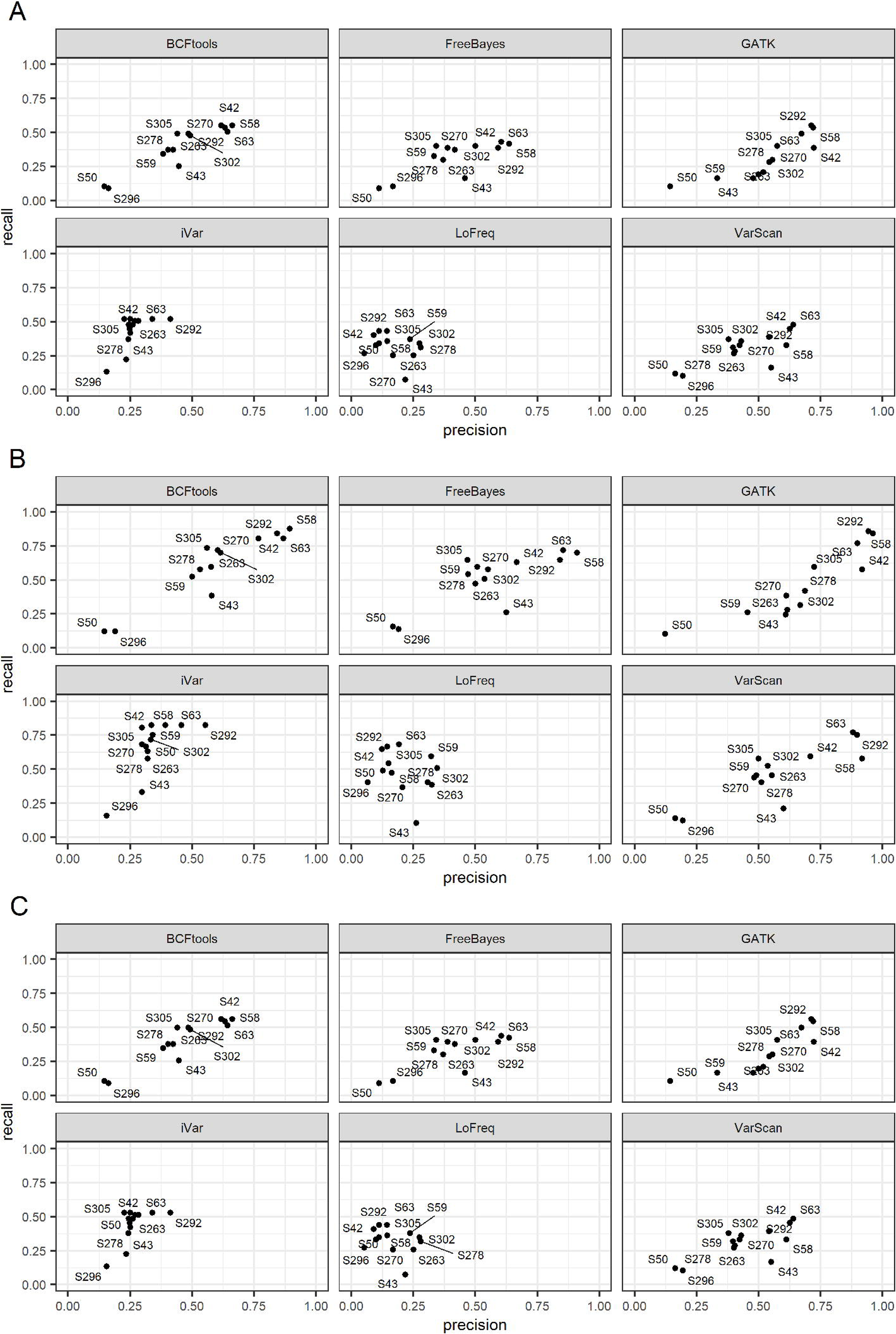
**A-C** Point plots of precision vs recall for real wastewater samples for the three Omicron variants, faceted by variant caller. A. Omicron England variant reference; B. Omicron Hong Kong variant reference; C. Omicron Australia variant reference. Since the wastewater samples are known to contain the Omicron variant, samples do show a higher precision and recall compared to the negative controls used to generate Figure 4A-B-D. Samples S63, S58, S42 and S292 had the highest precision and recall for BCFtools, Freebayes, GATK, iVar and VarScan, while LoFreq did not call with the same efficiency. S50 and S296 had the lowest precision and recall for all the callers. Notably, GATK did not call S296.

**Figure 4.**
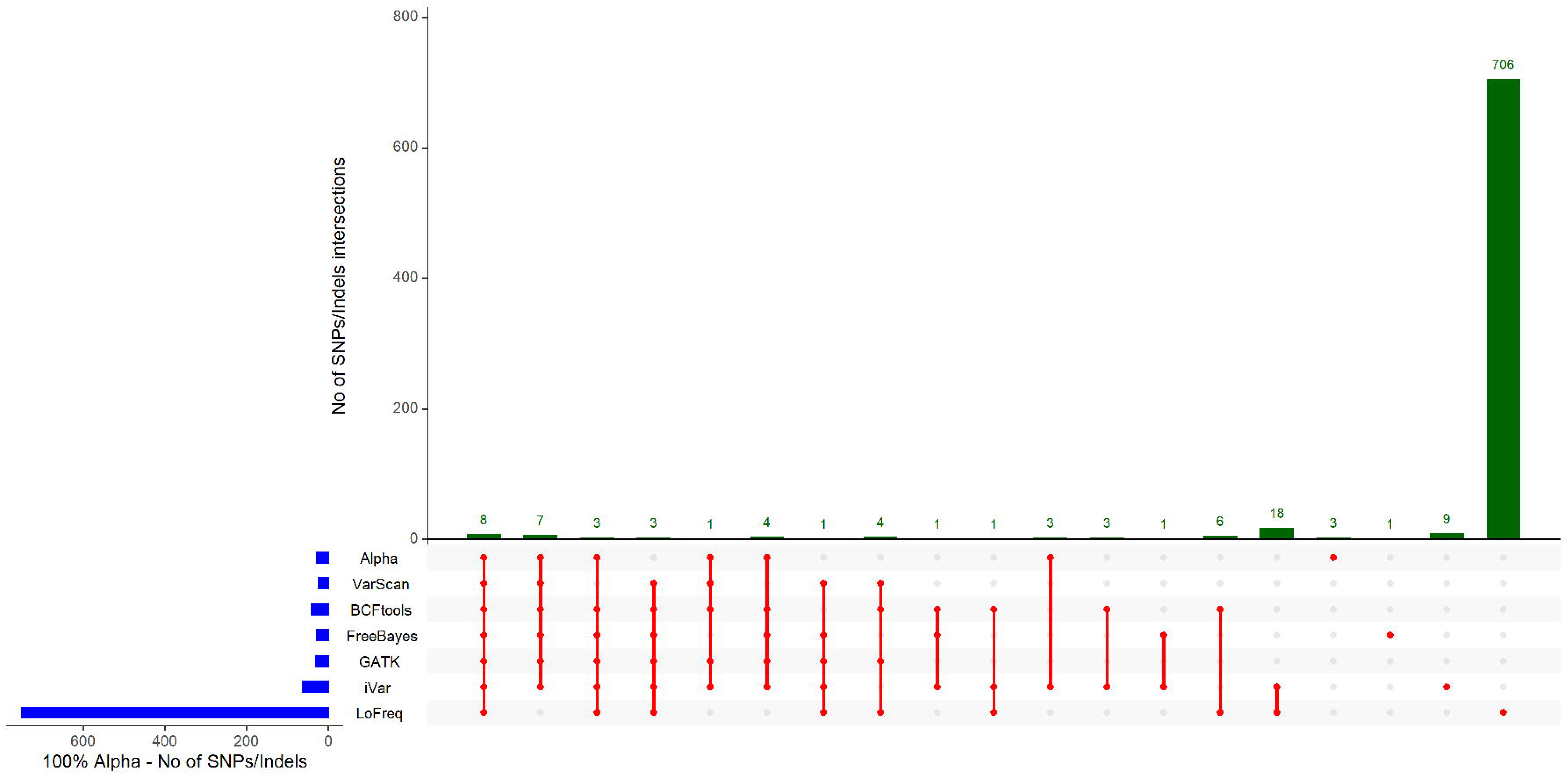
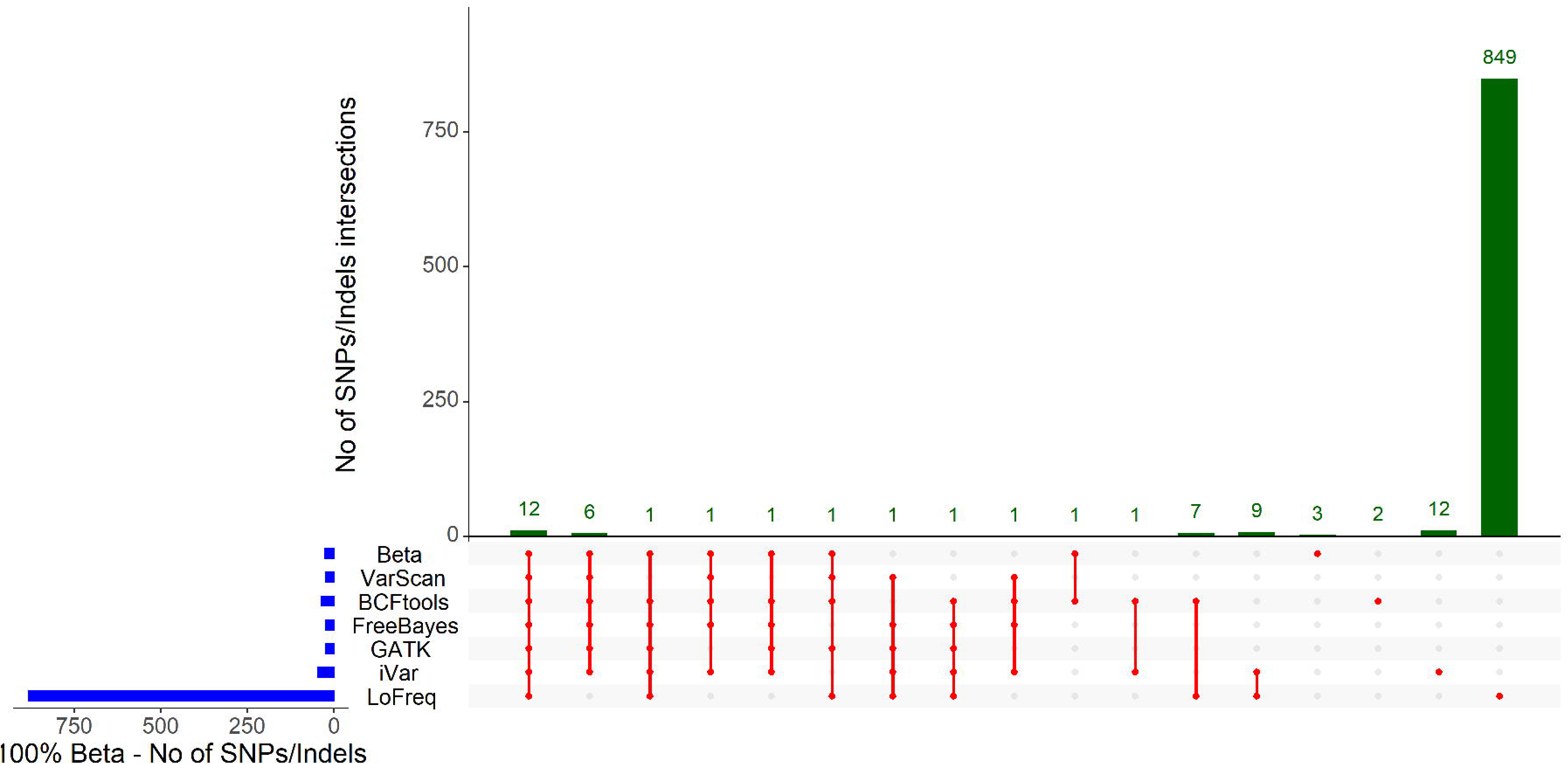
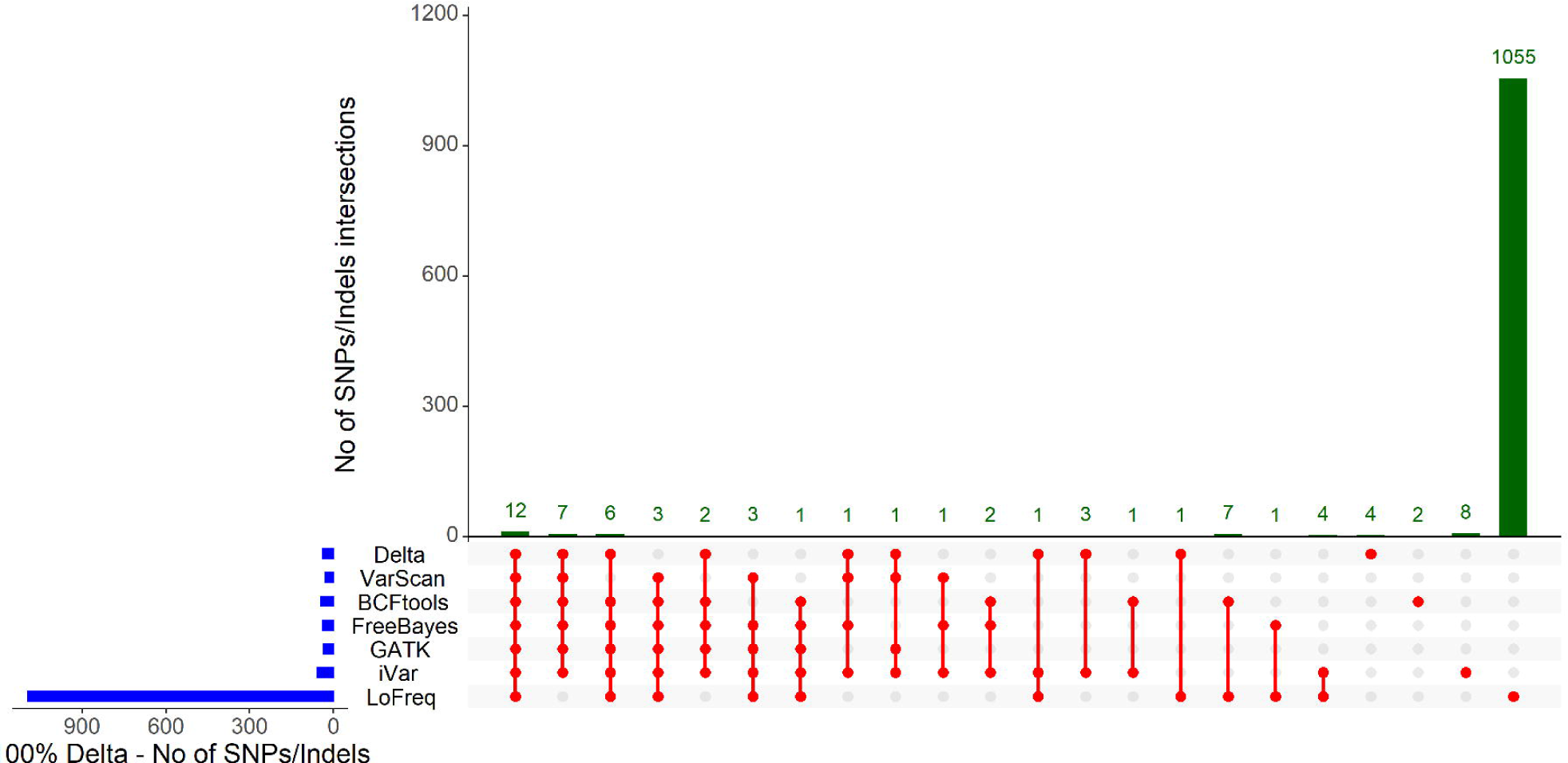
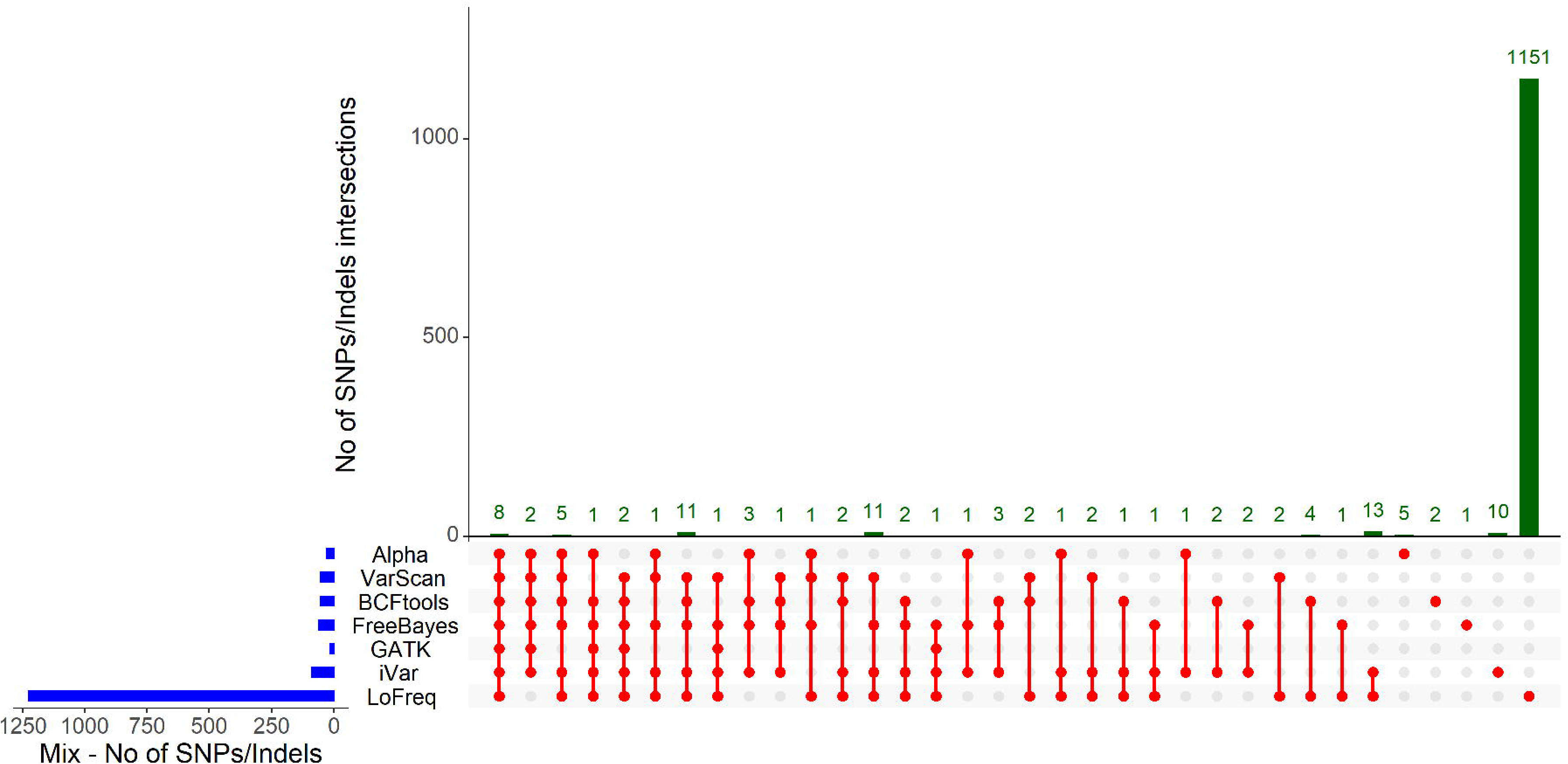
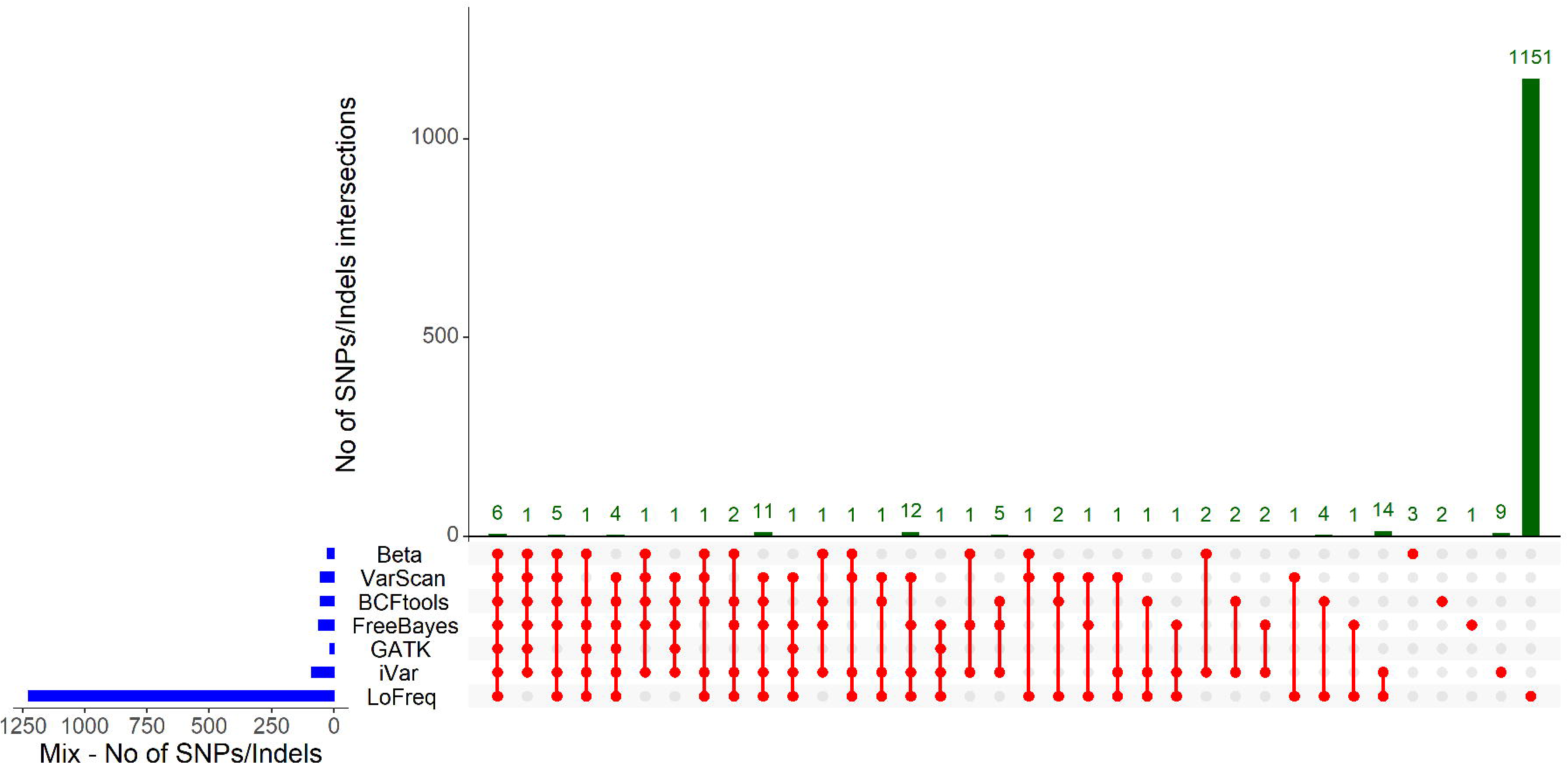
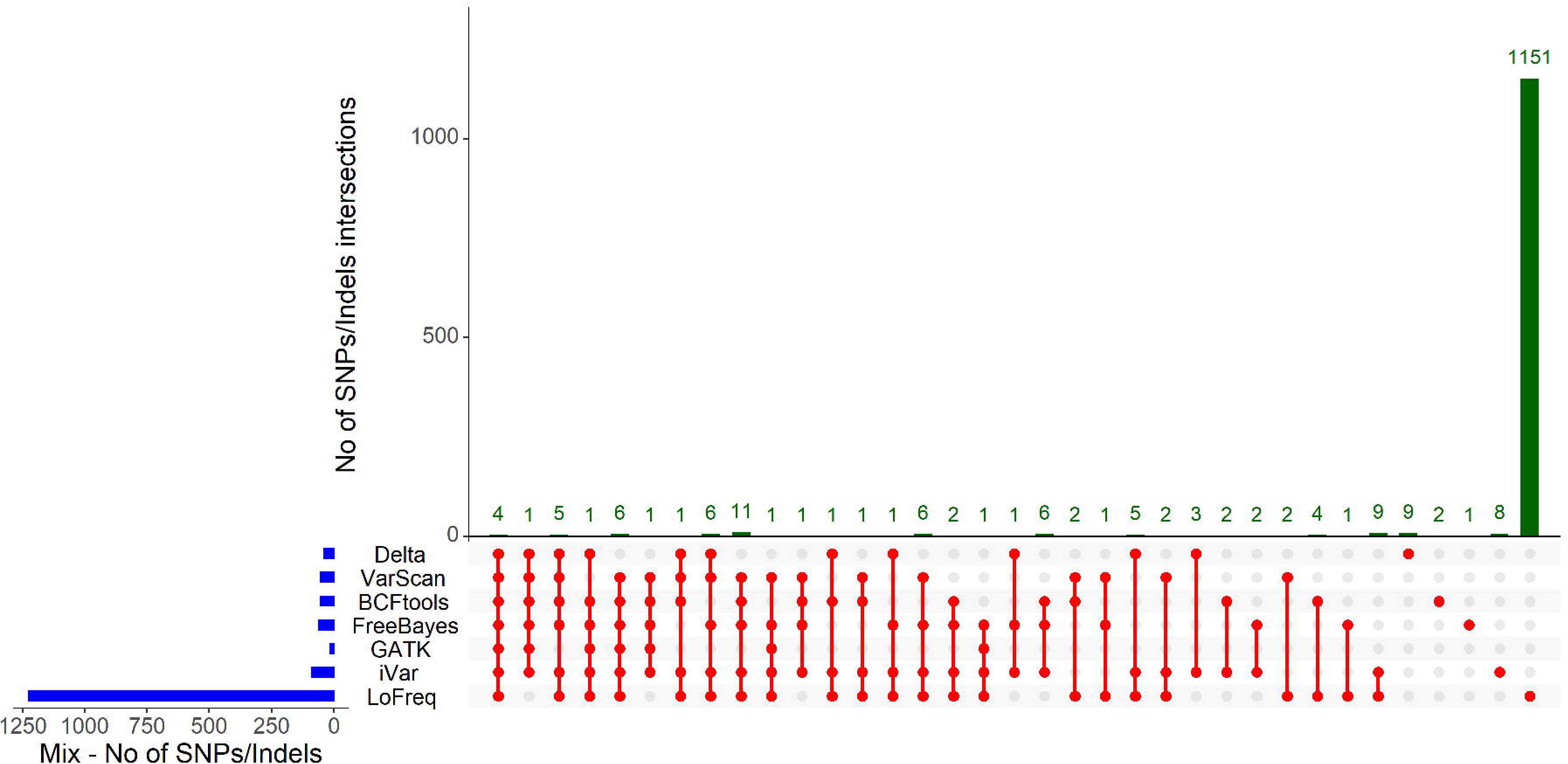
**A-F** Upset plots showing the common set of mutation found in (A) Alpha reference genome and sample containing 100% alpha synthetic genome (B) Beta reference genome and sample containing 100% beta synthetic genome (C) Delta reference genome and sample containing 100% delta synthetic genome. (D-F) Upset plots showing common mutation between sample mix (50% Alpha, 25% Beta and 25% Delta synthetic genome) and (D) Alpha reference genome (E) Beta reference genome and (F) Delta reference genome for 6 different variant callers. A= Alpha; B= Beta; C= Delta; D= mixed sample 12 Alpha; E= mixed sample 12 Beta; F= mixed sample 12 Delta. For each A-F plot we added the variant being tested at the top of the variant callers. Each mutation called by variant callers below Alpha, Beta and Delta can then be compared to look at how many of the variant-defining mutations are found by the variant caller. The Figures show that not all the defining mutations are found by each of the caller and the additional mutations each variant caller has found. Among all the callers, LoFreq is the caller with the highest number of mutations detected and much fewer corresponding to the variant-defining list as shown by the variant on top of the callers.

### Comparison of known variant defining mutations found in synthetic and wastewater samples across the six variant callers

We calculated the total number of known SNPs, Indels and MNPs (multi nucleotide polymorphisms) as described by Twist (synthetic samples) or PHE (wastewater samples) for each variant and compared these with those found in our synthetic (Figure 4A-F, Supplementary Figure 5A-F and Supplementary file 2) or wastewater samples (Figure 5A-F, Supplementary Figure 5A-F and Supplementary file 2) by each of the callers. SNPs/Indels bar plots were also shown in absence of LoFreq to show the divergence among all the callers on a different scale, as shown in Supplementary Figure 6A-D. In Figure 4A-F and 5A-F we used UpSet plots to show TPs for a subset of both synthetic and wastewater samples, respectively. For the synthetic samples we chose the three samples with 100% of a variant and one with a mix of the three (sample 12, Table 2). Among the wastewater samples, we chose six samples, representative of both Omicron lineages and Delta variants (Table 3, samples highlighted in bold). As shown in Figure 4A-F, and in concordance with the data presented above, LoFreq did call a much higher number of mutations compared to all the other variant callers in all the synthetic samples analysed, leading to a high number of FNs. When looking in more detail at how many of those were the defining mutations for each of the reference genomes (Table 1), we found that all callers identified the majority of the expected mutations for the variants being investigated, except LoFreq which only found 11/28 mutations for Alpha, 14/25 for Beta, and 20/37 for Delta (Figure 4A-B-C, Supplementary file 2). In addition, a set of 3 mutations was not detected by any of the variant callers for Alpha and Beta variants and 4 mutations for the Delta variant. The detailed number of defining expected mutations for all the callers are described in Supplementary file 2.

**Figure 5.**
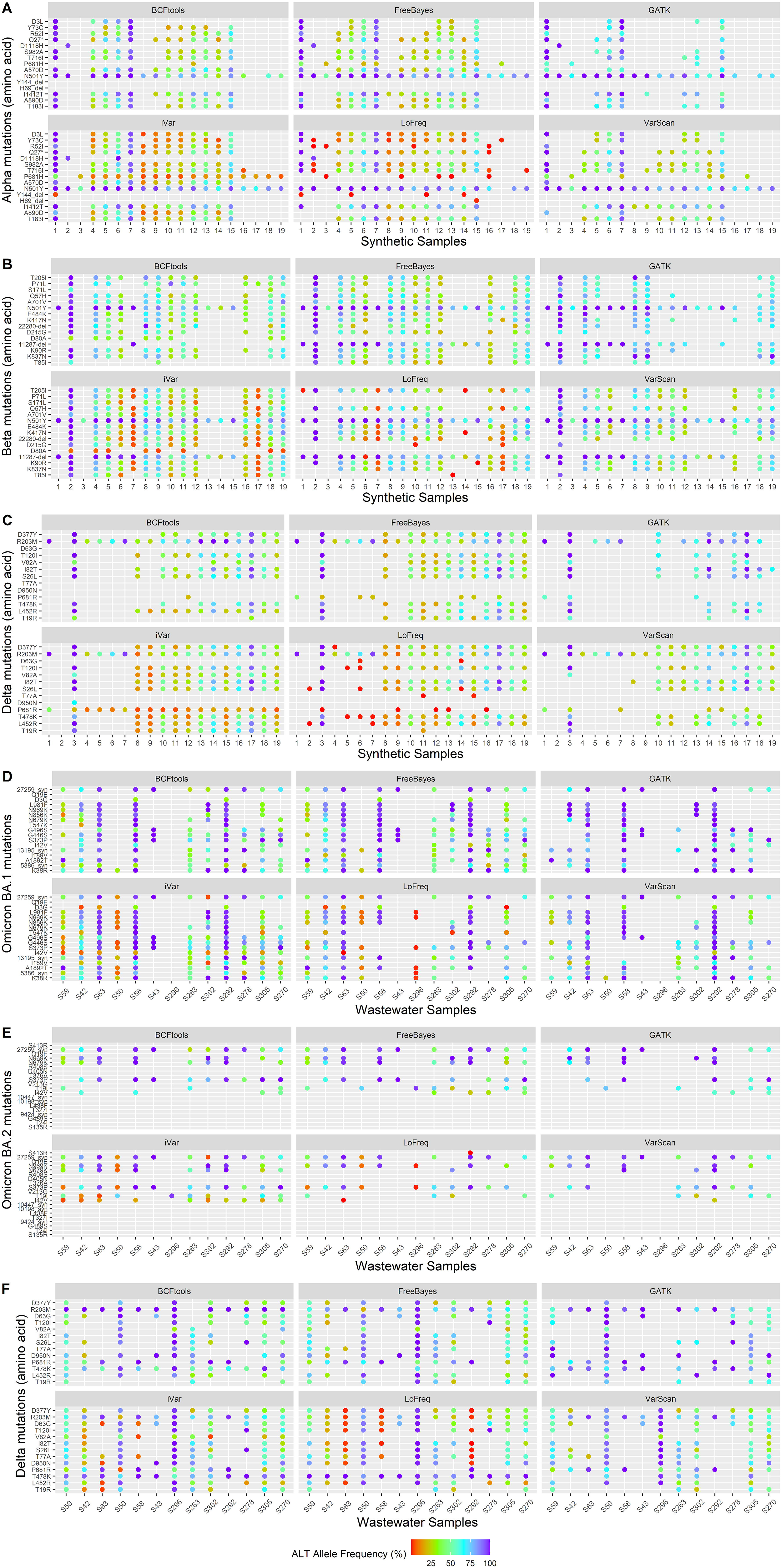
**A-F** Upset plots of wastewater samples showing the common set of defining mutations found between Delta, Omicron BA.1 and Omicron BA.2 and wastewater samples (A) S42 (B) S58 (C) S63 (D) S292 (E) 296 and (F) S263 for 6 different variant callers. A=S42, B=S58, C=S63, D=S292, E=S296 – GATK failed, F=S263. As described for Figure 4A-F, each mutation called by variant callers listed below Omicron BA.1, Omicron BA.2 and Delta can then be compared to look at how many of the variant-defining mutations are found by the variant caller. The Figure show that not all the defining mutations are found by each of the caller and the additional mutations each variant caller has found. As seen in the synthetic samples, LoFreq is the caller with the highest number of mutations detected and much fewer corresponding to the variant-defining list as shown by the variant on top of the callers. All callers also show the presence of unique mutations not found by the other callers and not present in the list of the defining ones as seen under Omicron BA.1, Omicron BA.2 and Delta.

For the synthetic mixed strain sample (Table 2, sample 12) we tested the presence of the defining mutations for Alpha, Beta and Delta (Figure 4D-E-F, respectively and Supplementary file 2) which were mixed in a 50:25:25 ratio, respectively. As summarised in Supplementary file 2, GATK called the lowest number of expected mutations for all the three variants, followed by VarScan and LoFreq. On the other hand, we found that iVar, FreeBayes and BCFtools were the callers with the highest number of expected mutations for all three variants profiled. Five mutations for the Alpha variants, 3 for the Beta and 8 for the Delta variant were not detected by any of the callers.

Similarly, we called variants for the wastewater samples (in bold Table 3, Figure 5A-F, Supplementary Figure 5B-D-F and Supplementary file 2) and observed the same pattern. More specifically, we investigated how many of the Delta, Omicron BA.1 and BA.2 defining mutations were detected by each of the variant callers across the samples and compared the numbers. As shown in Figure 5A-F, we found that, of all the mutations detected in the wastewater samples, 13/20 mutations for BA.2 were not detected by any of the variant callers (Supplementary file 2), yielding to a very low number of defining mutations found by each caller (up to 6 total mutations). For BA.1, who has a total of 17 defining mutations, LoFreq called the least number of expected mutations for sample S58 (7/17) and S263 (6/17). Overall, all the callers found between 6 to 16 defining mutations across the samples, with iVar having the highest number of expected mutations compared to the other callers: it detected all the 16/17 mutations for BA.1 in all samples, except 10/17 for S263 and 0/17 for S296. When looking at the Delta variant defining mutations, as observed for the other variants, there is a degree of difference within the same sample, e.g., iVar and LoFreq called 8/17 and 9/17 defining mutations, respectively for sample S63, but all the others only 3/17. Overall, iVar seemed to have performed well for the Delta variant where it called the highest number of expected mutations (Supplementary file 2).

Similarly, bar plots showing the number of total SNPs, Indels and MNPs across the samples for the six variant callers were also calculated. This is shown in Supplementary Figure 5A-F, for both synthetic and the real wastewater samples, and in absence of LoFreq in Supplementary Figure 6A-D. Interestingly, not all the variant callers were able to recognise MNPs. As shown in Supplementary Figure 5E-F only FreeBayes and iVar found this type of mutations across both synthetic and real wastewater samples.

### Comparison of alternate allele frequencies across the six variant callers

We extrapolated the alternate allele frequencies values from the vcf files for both the synthetic and wastewater samples across the six variant callers to look if these were called similarly. Degenerate codons were not plotted for any of the callers. In Figure 6A-C (synthetic) and Figure 6D-F (wastewater) we plot all the defining mutations for the variants of interest across all the 19 synthetic or the 13 wastewater samples. Frequencies are coloured by gradient.

**Figure 6.**
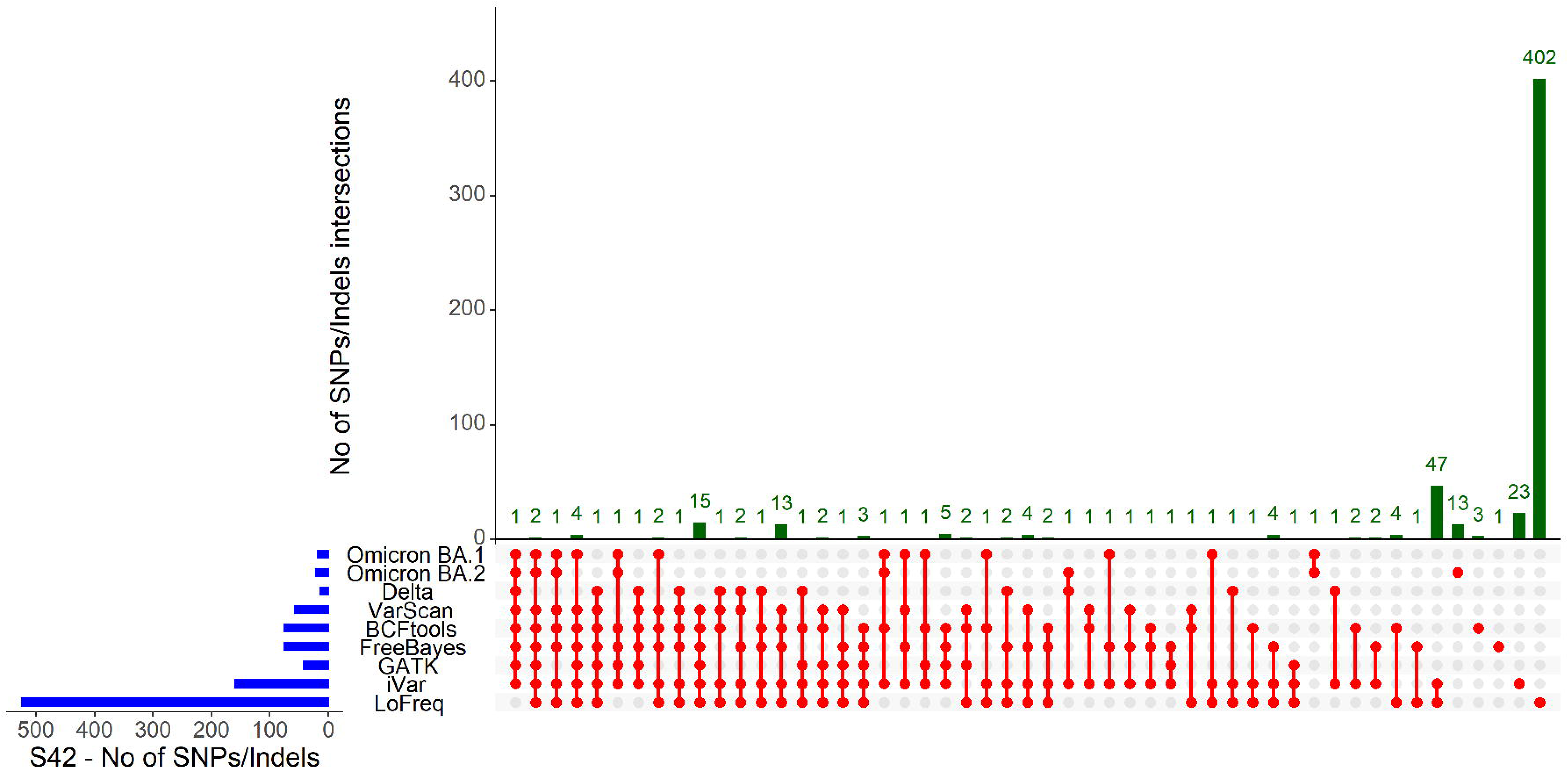
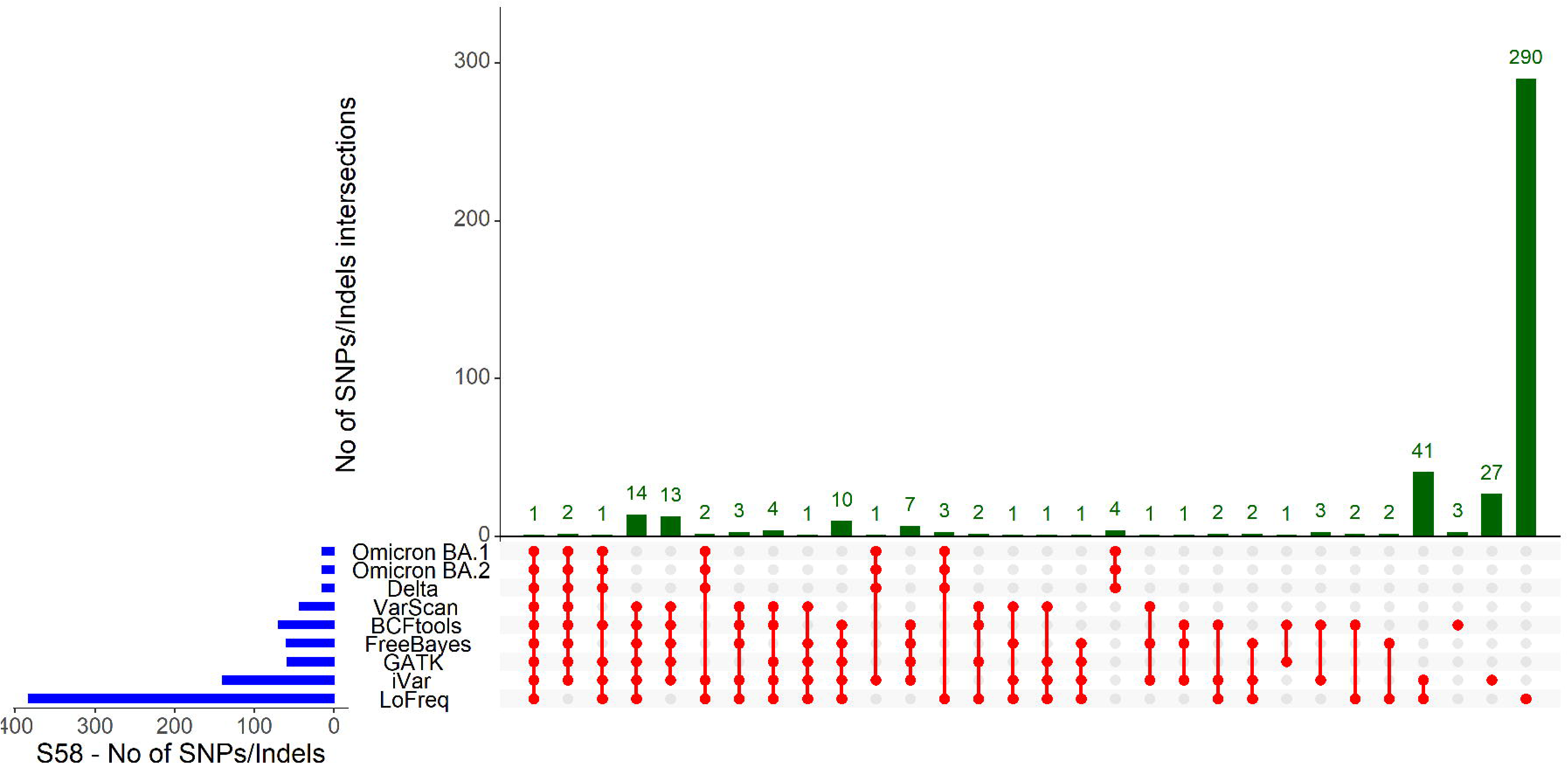
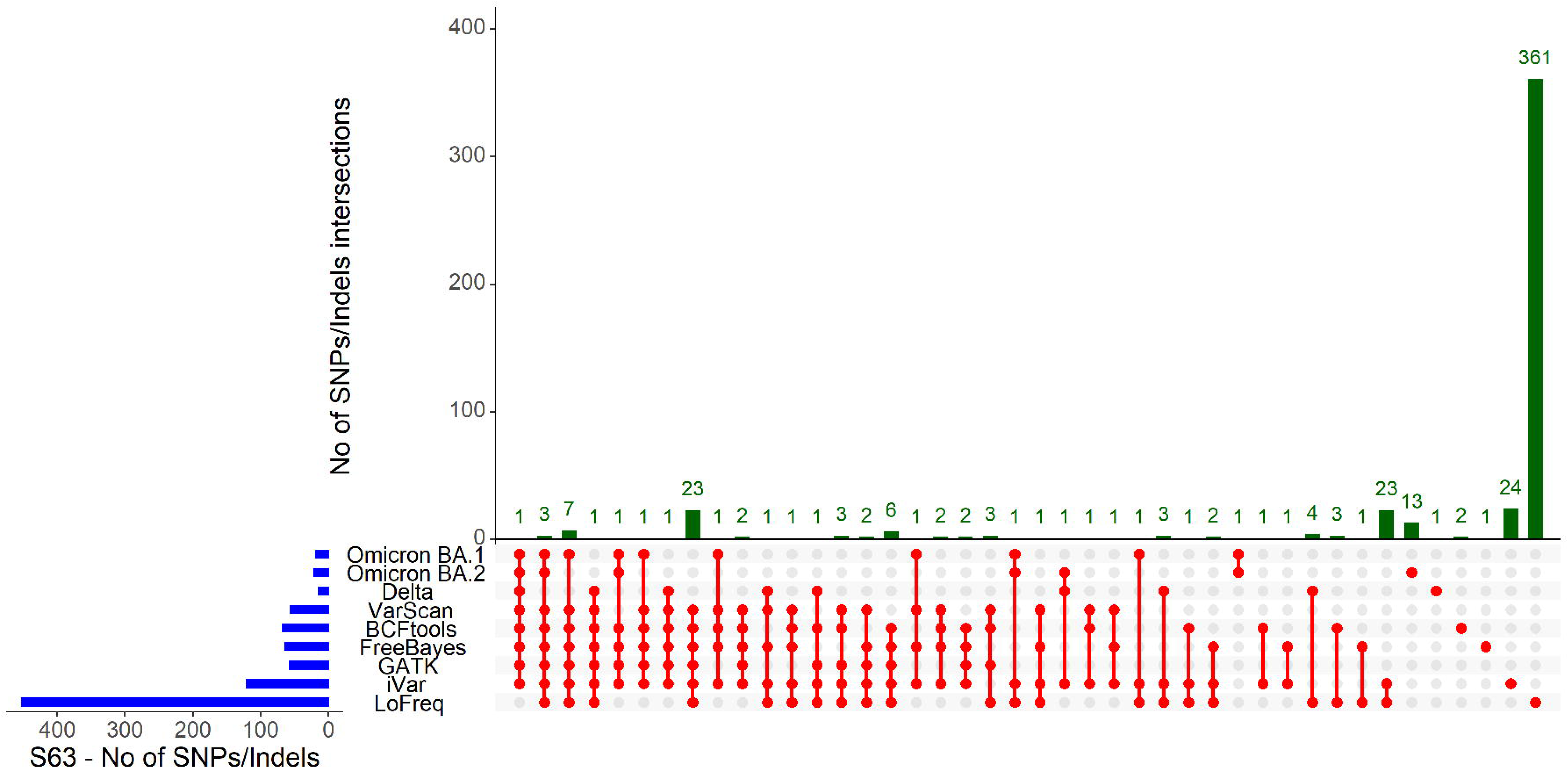
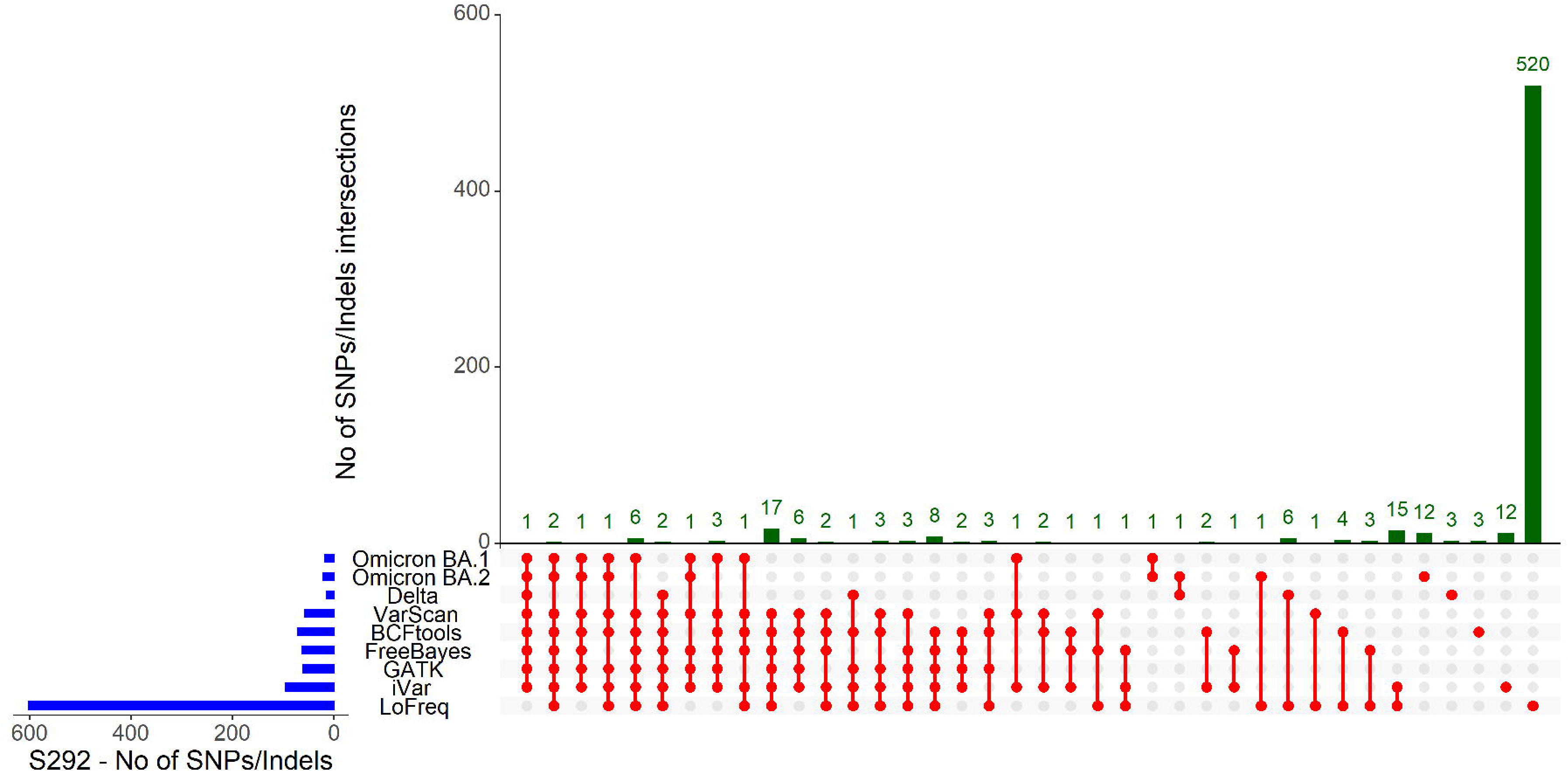
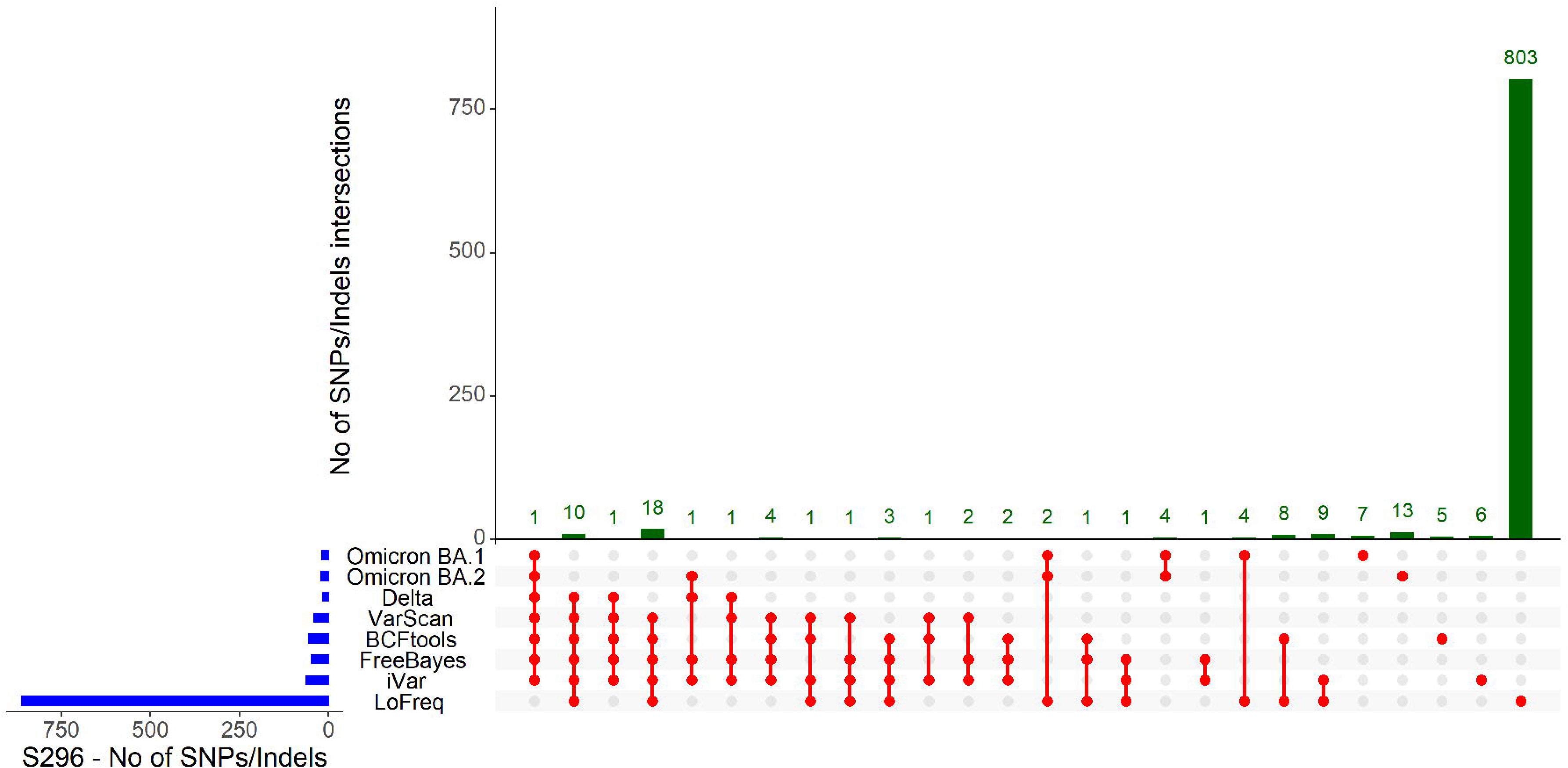
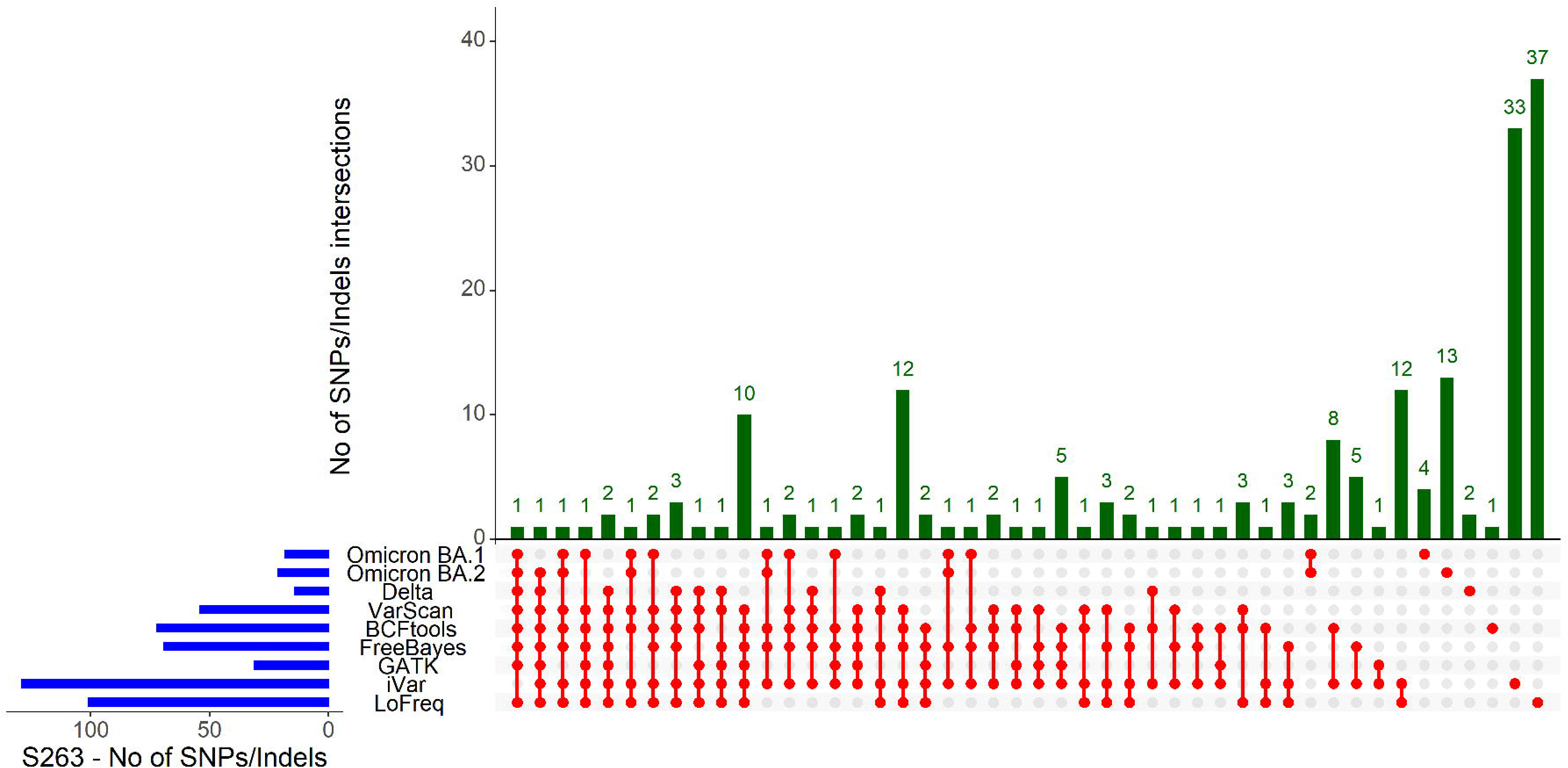
**A-F** Alternate allele frequencies of the defining mutations for Alpha (A), Beta (B) and Delta (C) variants were plotted for the 19 selected synthetic datasets for six different variant callers. Similarly, the alternate allele frequencies of the defining mutations of Omicron BA.1 (D), Omicron BA.2 (E) and Delta (F) were plotted for 13 wastewater samples for six different variant callers. The data points were coloured based on the 4-color rainbow gradient from red (0%) to purple (100%). Degenerate codons were not plotted.

Among the synthetic samples, all the callers had the same frequency for those samples where there was a high proportion of a variant (75-100%), such as samples 1 and 7 for Alpha (Figure 6A), sample 2 for Beta (Figure 6B) and samples 3 and 17 for Delta (Figure 6C). For the remaining samples, frequencies were similar, although iVar called more mutations than others, but at very low frequency. Similarly, for the wastewater samples we plotted frequencies for defining mutation for Omicron BA.1 (Figure 6D), BA.2 (6E) and Delta (6F) variants. BA.1 mutation frequencies were called at the same level across all the callers. In particular, samples S58, S63 and S292 were the ones with the highest number of mutations detected by all callers, at the same high frequency. All three samples were found to be positive for Omicron from previous data analysis (data not shown). Some mutations, such as Q19E were not called by any of the callers. It is worth highlighting that at the time the original samples were analysed (December 2021) there was no clear distinction between BA.1 and BA.2. Frequencies for BA.2 (Figure 6E) were also similar across the callers, although only a subset of these were detected, suggesting that more likely the BA.1 subgroup was the one circulating at that time. As for the Delta variant, two samples, S50 and S296, showed high frequency and were consistent among all the callers. Other samples were called similarly, with no evident differences.

## Discussion

In this paper we have analysed six variant callers commonly used in bioinformatics data analysis to empirically quantify their ability to identify mutations across mixtures of synthetic samples with known mutations as well as a set of wastewater samples with an unknown number of mutations. We first calculated recall and precision across the full genome for all the samples to define ground differences across the callers, to then focus in more detail on a set of defining mutations, by comparing how many of these were found and at which frequency by each of the callers. Our results suggest that the variant callers that showed the highest precision when looking at all samples together (synthetic and real wastewater samples) were GATK and VarScan followed by BCFtools and FreeBayes, which instead showed sparser data points. LoFreq and iVar showed the lowest precision values (Supplementary Figure 7C). Recall values were the lowest for GATK (and very sparse values), while they were significant statistical differences among the rest of the callers (in particular iVar’s stochastic dominance). Overall, the F1 score confirmed that LoFreq was the least sensitive (Supplementary Figure 7C), presenting numbers of mutations that are magnitudes larger than the rest, and subsequently with a lower precision. On the other hand, when focusing on selected mutations, iVar identified the highest number of expected defining mutations across both synthetic and wastewater samples, compared to the other callers.

Wastewater-based epidemiology (WBE) has been used for many years to monitor key pathogens such as polio [41-45]. However, it has undergone a renaissance during the SARS-CoV-2 pandemic, with many tools and software specifically designed to detect the virus in wastewater and being developed in the wake of WBE monitoring [46-50]. As such, tools used to detect the virus in clinical samples were used as templates, but in many cases, these did not reflect the composition of the wastewater samples accurately, e.g., the mixed strain nature as well as degradation of the viral RNA in the environment, thus the lack of a complete genome, and more importantly the lack of a consensus sequence. These nuances impact and should inform downstream analysis at the bioinformatics level: sequences could only be a fraction of the genome, as the samples may be highly degraded, and the lack of a consensus will affect the ability to assign a variant to a sample [51, 52]. Subsequently variant analysis is limited to a shorter region in some cases and variant assignment has to happen based on specific mutations known to define a variant, as used for clinical cases [53]. For this reason, a variant caller that adapts to the type of data available is very important as it will be expected to call the mutations with higher sensitivity.

The six callers analysed in this paper have many aspects in common as well as significant differences. For example, of all the callers LoFreq is the only one that requires base quality information to call variants, making this method much more stringent and robust than the others, but similarly prone to call many more mutations than expected [21], as seen in our results. In addition, it has been reported that LoFreq can efficiently recognise sequencing errors from expected mutations in non-environmental samples, however, it seems that it did not behave as efficiently in our wastewater samples nor with the synthetic samples. This could also be a consequence that more specific combinations of parameters have to be chosen for efficient performance. Although for some samples the recall was correct, the precision was lower indicating the lower efficiency. Indeed, LoFreq is a fast and sensitive variant caller that calls many mutations that, however, are not true positive, hence low recall and precision as found in our results. We suggest that this tool is more suited for shorter viral sequences, and more likely in samples with higher coverage. Similarly, GATK was designed to detect genomes across a range of sample sources, but not environmental, and this is reflected in the highly comprehensive set of parameters available to efficiently analyse the datasets (over 111 parameters) [20], but most of GATK parameters are not applicable to wastewater datasets. These aspects also highlight the difficulty in using these two tools for wastewater data, in contrast to FreeBayes, BCFtools or VarScan, which are functional and easy to apply in non-clinical settings. Nevertheless, we found that compared to most of the callers analysed, GATK did find the majority of the expected mutations (TPs) as well as overall good scores for recall and precision, suggesting that applying different sets of parameters might improve its functionality for environmental samples as well. This applies to LoFreq as well.

The selected variant callers have been extensively used in other fields for comparison purposes. For example, a recent paper comparing the efficiency of different mappers and callers in plant NGS data found that GATK was the best caller among those tested, suggesting that the type of data greatly affects not only the results but also the choice of tools used to analyse the datasets [30, 54, 55]. Although GATK was not the best of the callers in our study, we suggest these results are valid given the diversity of the datasets; namely, the plant genome being of better quality and not containing mixed strains compared to the wastewater, secondly, the fact that not always the expected mutations are known, thus many more mutations will account as TP or FP. This is independent of the pathogen studied, since most of the tools are widely applicable in different fields. Similarly, another study looking at exome sequencing also found that among the variant callers analysed, GATK UnifiedGenotyper performed best [54]. We suggest that looking at shorter regions of a genome, such as exons, has its advantage since it allows us to work with a relatively smaller and highly covered region. In addition, compared to the above paper, we used GATK Haplotypecaller, which is known to have a different algorithm for calling variants than GATK UnifiedGenotyper.

It should also be noted that many of the parameters within each of these variant callers were left in default in our analysis. In fact, the wide choice of parameters poses a risk on its own when comparing different tools as it introduces biases. In an ideal setting, the correct procedure will imply that all parameters are tested and those reflecting an outcome that is expected are then chosen. In this paper we did not assume a certain output as we did not use all the possible parameters and because we were not expecting similar results between the callers. Indeed, a small test using FreeBayes showed us that changing certain key parameters yielded many different outputs, all of them being acceptable results (data not shown). Because these parameters are not shared or found in other callers, the comparison could not be achieved, as it would have introduced an advantage or disadvantage for some callers. This is in agreement with current literature, where on many occasions’ parameters are left in default [30-32]. A direct consequence of this is that many results across our data would have had a different outcome. Indeed, LoFreq results might differ enormously if we had considered and adjusted all the parameters accordingly, irrespective of whether these were common to other variant callers.

It should be noted that calculating recall and precision for samples containing a mix of variants (two or more), is a cumbersome task, as some mutations can be shared among the variants, which will affect the ground truth. At the time of writing Deng et al., [35] designed a tool, where only samples with a mix of two genomes can be used. However, as of now there is no tool available for samples with a mix of three or more genomes. Wastewater are mixed samples, sometimes containing more than two variants. However, this notion will only be confirmed overtime, through sequencing of clinical cases. Therefore, with the aim to reflect real wastewater data, we calculated recall and precision for each variant known to be prevalent as it would be at that time that specific variant would have been circulating. This will inevitably overestimate the number of FP in each run as it should be calculated only once per sample, but it is however expected: any position not found to be a TP for one variant, will show as a FP. But, by testing each of the variants separately, this will give us the correct TP which are the values we have been using in our manuscript to compare the callers (defining mutations).

## Conclusion

In conclusion, we suggest that callers such as Varscan, BCFtools and Freebayes are overall preferable (Supplementary Figure 7C), particularly when mutations are not known as they are called with higher specificity and sensitivity.

However, if specific mutations are under investigation and expected in the output, such as the ones we used as variant-defining, iVar performed best.

We also suggest that, upon choice of one variant caller for a specific study other than comparison purposes, all parameters should be explored and tested to better improve the calling capability.

In the future, tools that can analyse mixed samples without the need to run the strains separately are also preferred as they will give even more accurate values of recall and precision.

## Author contribution

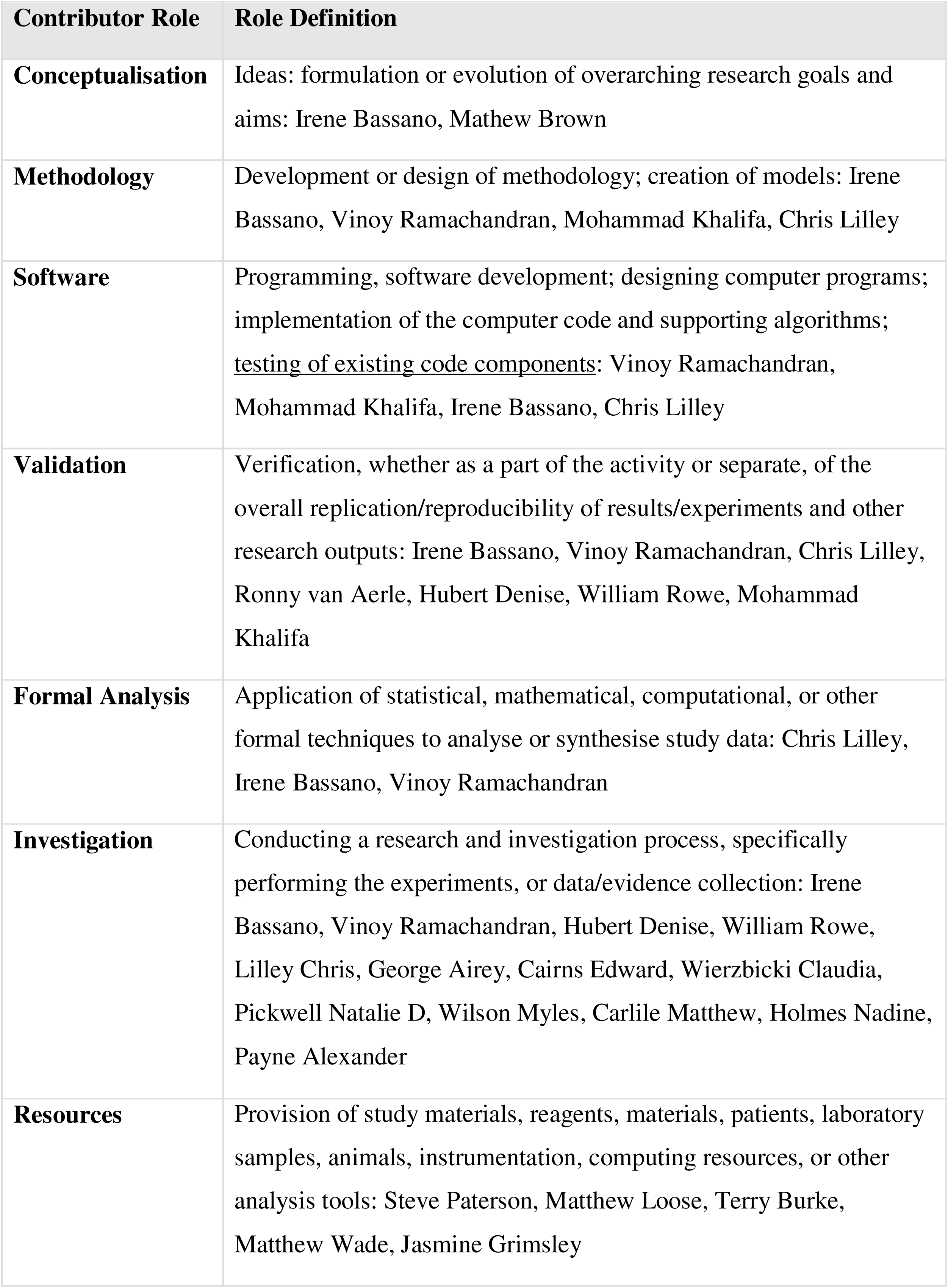
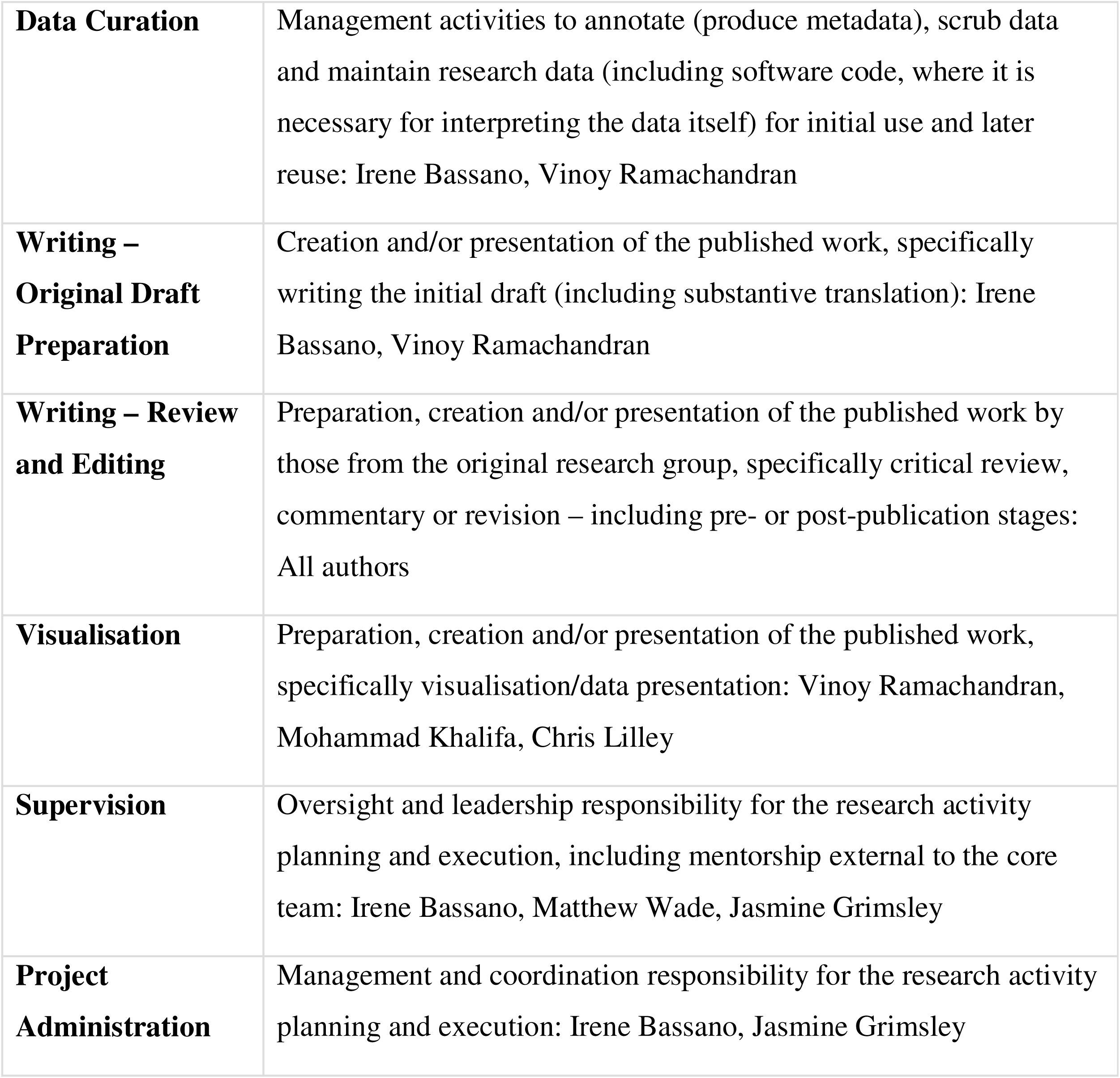

## Data Availability

All data produced in the present study are available upon reasonable request to the authors

## Acknowledgements

We are grateful to Dr Zhi-Luo Deng from the Department of Computational Biology of Infection Research, Helmholtz Centre for Infection Research for helpful discussions and comments, help in installing, running and updating scripts to calculate recall and precision (Quasimodo), Rachel Tucker and Tom Holden from NERC Environmental Omics Facility, Ecology and Evolutionary Biology, School of Biosciences, University of Sheffield for technical assistance.

## Data availability

Sequencing data is available upon request.

## Figures and Tables legends

**Supplementary Figure 1 A-D** Point plots of precision vs recall for synthetic samples, grouped and coloured by variant caller and a linear regression for each. A, Alpha variant reference, B. Beta variant reference, C. Delta variant reference and D. Gamma variant reference. The figure shows the low precision recorded by LoFreq compared to the rest of the variant callers while is higher for VarScan, FreeBayes, BCFtools and GATK. Interestingly, trend lines overlap for Freebayes and BCFtools.

**Supplementary Figure 2** Plot of F1 score vs callers for three synthetic samples (1) 100% alpha (2) 100% Beta and (3) 100% Delta as listed in Table 2 (samples with *). Each plot represents caller in relation to either the precision or the recall, expressed as the F1 score. The figure shows that all the replicates (filled circle) have similar recall and precision, suggesting that no major differences are observed. As shown in Figure 1 and 2, LoFreq shows the lowest F1 score compared to the rest of the callers, followed by iVar. FreeBayes and VarScan, showed similar results, while GATK had the highest F1 score, followed by BCFtools.

**Supplementary Figure 3 A-D** Point plots of precision vs recall for wastewater samples, coloured by variant caller. A, Alpha variant reference, B. Beta variant reference, C. Delta variant reference and D. Gamma variant reference. The figure shows that variants not found in the mix such as Alpha, Beta and Gamma have low precision and recall. Some of the samples are known to be positive for the Delta variant therefore the latter will have a higher precision and/or recall.

**Supplementary Figure 4 A-C** Point plots of precision vs recall for wastewater samples for the Omicron variant, grouped and coloured by variant caller and a linear regression for each. A, Omicron England variant reference, B. Omicron Hong Kong variant reference, C. Omicron Australia variant reference. Since the wastewater samples are known to contain the Omicron variant, samples do show a higher precision and recall compared to the negative controls used to generate Figure 4 A-B-D.

**Supplementary Figure 5 A-F** SNPs, Indels, MNVs bar plots for synthetic and real wastewater samples.

A. Number of SNPs calculated for each of the 19 synthetic samples (mean of replicates). The figure clearly identifies LoFreq as the caller with the highest number of SNPs detected, while the rest of the callers do show a similar pattern. Detailed differences excluding LoFreq can be appreciated in Supplementary Figure 2A-B.

B. Number of SNPs calculated for each of the 13 wastewater samples with variable results among the callers. In comparison to other callers, LoFreq still calls more SNPs than expected in some of the samples, namely S296, S292, S42, S50.

C. Number of Indels calculated for each of the 19 synthetic samples (mean of replicates). As seen for the SNPs bar plots, LoFreq calls the highest number of Indels detected, while the rest of the callers do show a similar pattern. Detailed differences excluding LoFreq can be appreciated in Supplementary Figure 2C-D.

D. Number of Indels calculated for each of the 13 wastewater samples with variable results among the callers. In comparison to other callers, LoFreq still calls more Indels than expected in some of the samples, namely S296, S292, S305, S42, S50, S58 and S63. Notably, we were not able to verify the presence of Indels for GATK for sample S296.

E. Number of MNPs calculated for each of the 19 synthetic samples (mean of replicates). Only Freebayes and iVAR had detectable values to be plotted, while the rest of the callers did not call this type of base variation.

F. Number of MNPs calculated for each of the 13 wastewater samples. As seen for the synthetic samples, only Freebayes and iVAR detected the presence of MNPs.

**Supplementary Figure 6 A-F** SNPs, Indels, MNVs bar plots for synthetic and real wastewater samples without plotting LoFreq values. A. Number of SNPs calculated for each of the 19 synthetic samples (mean of replicates). B. Number of SNPs calculated for each of the 13 wastewater samples. C. Number of Indels calculated for each of the 19 synthetic samples (mean of replicates). D. Number of Indels calculated for each of the 13 wastewater samples. E. Number of MNPs calculated for each of the 19 synthetic samples (mean of replicates). F. Number of MNPs calculated for each of the 13 wastewater samples.

**Supplementary Figure 7 A-C** Statistical analysis of recall, precision and F1 score across the six variant callers for synthetic and wastewater samples. The figure shows boxplots showing variant caller scores for wastewater samples. For each figure, Top: Precision. Middle: Recall. Bottom: F1 Score. A. Boxplots showing variant caller scores for synthetic samples. Right: Post-hoc Dunn’s test p-values, highlighted where p < 0.05 indicating significant difference between the distribution of scores of that caller with another. LoFreq is stochastically dominated by the others when evaluating precision and F1 scores. iVar is stochastically dominant for recall. B. Left: Boxplots showing variant caller scores for wastewater samples. Right: Post-hoc Dunn’s test p-values, highlighted where p < 0.05 indicating significant difference between the distribution of scores of that caller with another. LoFreq is stochastically dominated by the others when evaluating precision and F1 scores. iVar is stochastically dominant for recall but is dominated by all callers except LoFreq for precision.

C. Left: Boxplots showing variant caller scores for synthetic and wastewater samples combined. Right: Post-hoc Dunn’s test p-values, highlighted where p < 0.05 indicating significant difference between the distribution of scores of that caller with another. LoFreq is stochastically dominated by the others when evaluating precision and F1 scores. iVar is stochastically dominant for recall but is dominated by all callers except LoFreq for precision.

