## Supplementary file 1 for "Evaluation of variant calling algorithms for wastewater-based epidemiology using mixed populations of SARS-CoV-2 variants in synthetic and wastewater samples"

Supplementary Information – commands and parameters used

**Commands**

**Index the sorted bam file:**

samtools index sample1.sorted.bam

**bcftools:**

bcftools mpileup -Ou -f ref.fa sample1.sorted.bam| bcftools call -mv -Ob -o sample1.bcf

bcftools convert -O v -o sample1.vcf sample1.bcf

**Freebayes:**

freebayes -f ref.fa sample1.sorted.bam > sample1.vcf

freebayes -f ref.fa --min-coverage 8 -F 0.01 -q 15 -P 0.05 sample1.sorted.bam > sample1-mod.vcf

**iVar:**

samtools mpileup -aa -A -d 1000000 -B -Q 0 sample1.sorted.bam | ivar variants -p sample1-ivar.tsv –q 20 -t 0.03 -r ref.fa

#iVar generates tsv file. This python script converts tsv to vcf format

python3 ivar_tsv_to_vcf.py $ sample1-ivar.tsv $ sample1-ivar.vcf

**lofreq:**

To add indel quality score into bam file

lofreq indelqual -f ref.fa --dindel -o sample-indel.bam sample.bam

To run

lofreq call-parallel --pp-threads 10 --call-indels -f ref.fa -o sample-indel.vcf sample-indel.bam

**varscan:**

samtools mpileup -f ref.fa -q 10 -d 1000000 sample1.sorted.bam > sample1.pileup

cat sample1.pileup | awk 'BEGIN {FS="\t";OFS="\t";print "#chr","position","refbase","coverage"} {print $1,$2,$3,$4}' > sample1.cov.tsv

varscan pileup2snp sample1.pileup --p-value 0.05 > sample1.varscan.tsv

sequenza-utils pileup2acgt -p sample1.pileup > sample1.acgt.tsv

varscan pileup2indel sample1.pileup --p-value 0.05 > sample1.varscan.indel.tsv

varscan mpileup2snp sample1.pileup --p-value 0.05--output-vcf 1 > sample1-snps.vcf

varscan mpileup2indel sample1.pileup --p-value 0.05 --output-vcf 1 > sample1-indel.vcf

vcfcat sample1-snps.vcf sample1-indel.vcf > sample1.vcf

bgzip sample1-snps.vcf

bgzip sample1-indel.vcf

bcftools index sample1-snps.vcf.gz

bcftools index sample1-indel.vcf.gz

bcftools concat sample1-snps.vcf.gz sample1-indel.vcf.gz -o sample1-merged.vcf

**GATK Haplotypecaller:**

picard AddOrReplaceReadGroups I=sample.bam O=sample-dedup.bam RGID=4 RGLB=lib1 RGPL=ILLUMINA RGPU=unit1 RGSM=20

gatk --java-options "-Xmx4g" MarkDuplicatesSpark -I sample-dedup.bam -O sample-dedup-Markdup.bam

gatk --java-options "-Xmx4g" HaplotypeCaller -R ref.fa -I sample-dedup-Markdup.bam -O sample.vcf

### Quasimodo:

<u>To run quasimodo with 4 or less vcf files</u>

python3 run_benchmark.py vareval -t 20 -c ~/miniconda3/envs -v sample2.vcf,sample2.vcf, sample3.vcf, sample4.vcf -r ref.fa,variant.fa -o OUTPUT_DIR

<u>To run quasimodo with more than 4 files</u>
python3 run_benchmark.py vareval --novenn -t 20 -c ~/miniconda3/envs -v sample2.vcf,sample2.vcf,sample3.vcf,sample4.vcf……samplen.vcf -r ref.fa,variant.fa -o OUTPUT_DIR

**Frequency Plots:**

**Commands used to extract values from vcf files.**

**BCFtools**

bcftools query -f '%POS\t%INFO/DP\t%INFO/DP4\n' vcf_file_name

**VarScan**

bcftools query -f '%POS\t%FORMAT/FREQ\n' vcf_file_name

**iVar**

bcftools query -f '%POS\t%FORMAT/ALT_FREQ\n' vcf_file_name

**LoFreq**

bcftools query -f '%POS\t%INFO/AF\n' vcf_file_name

**FreeBayes**

bcftools query -f '%POS\t%INFO/DP\t%INFO/DPB\t%INFO/SRF\t%INFO/SRR\t%INFO/SAF\t%INFO/SAR\n' vcf_file_name

**GATK**

bcftools query -f '%POS\t%FORMAT\n' vcf_file_name
